# Islet-specific DNA hypomethylation identifies gene enhancer loci driving Type 2 Diabetes risk

**DOI:** 10.1101/2025.09.09.25335336

**Authors:** Verda Miranda, Timothy J. Scott, Elizabeth Dorans, Shaojun Mei, Adam X. Miranda, Al Powers, Lea Davis, Marcela Brissova, Emily Hodges

## Abstract

Genome-wide association studies (GWAS) have identified hundreds of genetic loci linked to pancreatic islet dysfunction and type 2 diabetes (T2D). Many of these loci map to islet genes and their enhancers; however, a complete understanding of the gene regulatory contributions to T2D is lacking. We have previously shown that cell-type specific DNA hypomethylation patterns identify gene enhancers that contribute to the heritability of cell-type relevant traits. Here, we determine a reference dataset of islet hypomethylated regions (HMRs) from DNA methylomes of donors without T2D. Comparing islet HMRs to profiles from functionally diverse cell types, we identify ∼4,800 islet-specific HMRs and demonstrate their enrichment for key genes and transcription factor motifs essential for β cell function. We further show that islet-specific HMRs are highly enriched for GWAS variants that contribute specifically to T2D heritability. Leveraging genotype-phenotype data from the Vanderbilt BioVU biobank, which links patient DNA to electronic health records (EHR), we perform phenome- and lab-wide association scans that replicate islet HMR associations with T2D and uncover novel links to related clinical traits. To overcome challenges posed by stringent GWAS significance thresholds, we develop an HMR-WAS, which allows detection of T2D signals in our smaller BioVU cohort. Our comprehensive framework reveals enhancers of genes involved in insulin secretion and glucose metabolism and demonstrates the impact of T2D-associated variants on their transcriptional activity. By integrating HMRs with patient EHR, our work underscores the potential to reveal new enhancer loci associated with disease risk, especially those missed in a conventional GWAS.

## BACKGROUND

A complex interplay exists between genetic and biological processes that shape Type 2 Diabetes (T2D). Over the past decade, genome-wide association studies (GWAS) have collectively identified genetic markers of T2D risk, with the largest multi-ancestry GWAS to date revealing 611 risk loci representing 1,289 independent association signals (p-value < 5.0 × 10^−8^)(Suzuki et al. 2024). Despite these significant findings, T2D-associated variants do not completely explain the heritability of T2D, with recent heritability estimates varying between 25-80%(Poulsen et al. 1999; Armstrong et al. 2024; Kaprio et al. 1992; Almgren et al. 2011; Manolio et al. 2009). Furthermore, T2D is a complex, polygenic trait that is likely driven by a network of variants with small effect sizes. These true positive associations are often overlooked, even in well-powered studies. Therefore, strategies that uncover the “missing biology” underlying T2D are needed.

Integration of T2D GWAS and epigenomic data from human islets have enabled functional annotation of single nucleotide polymorphisms (SNPs), revealing that SNPs associated with T2D and related traits frequently occur within islet gene enhancers(Gaulton et al. 2015; Pasquali et al. 2014; Parker et al. 2013; Varshney et al. 2017; Stitzel et al. 2010; Greenwald et al. 2019; Cebola 2019; Thurner et al. 2018). These and other studies argue that the majority of T2D risk variants influence disease susceptibility by disrupting gene expression and regulatory modules in β cells(Pasquali et al. 2014; Parker et al. 2013; Varshney et al. 2017; Walker et al. 2023; Thurner et al. 2018; Chiou et al. 2021a; Rosengren et al. 2012). Studies using animal and cellular models have demonstrated that T2D risk variants alter enhancer activities through changes in local chromatin accessibility, target gene expression, and/or transcription factor (TF) binding affinity. Additionally, these regulatory changes were associated with impaired insulin secretion and reduced islet β cell mass(Parker et al. 2013; Pasquali et al. 2014; Gaulton et al. 2015; Olsson et al. 2014; Varshney et al. 2017; Dayeh et al. 2014; Scott et al. 2017; Keaton et al. 2016; Florez 2008; Scott et al. 2012; Szczerbinska et al. 2022; Dupuis et al. 2010). Thus, the application of epigenomic data to GWAS findings has unveiled islet regulatory mechanisms underlying T2D susceptibility. Despite progress in this area, the full scope of islet gene regulatory contributions to T2D remains unknown.

A better understanding of the gene regulatory mechanisms driving T2D requires comprehensive identification of gene enhancers critical to cell identity and function, which we have shown is achievable through comparative DNA methylation profiling. Whole genome methylation studies from our lab and others have demonstrated that hypomethylated regions (HMRs) form discrete patterns associated with gene regulatory elements, including promoters and enhancers(Gu et al. 2016; Schlesinger et al. 2013; Roadmap Epigenomics Consortium et al. 2015; Neri et al. 2017). By comparing the HMR profiles of distinct cell types and developmental stages, we showed that cell-type and lineage-specific HMRs correspond to key lineage-determining enhancers(Gama-Sosa et al. 1983; Meissner et al. 2008; Portela and Esteller 2010; Hodges et al. 2011; Schlesinger et al. 2013; Molaro et al. 2011; Bock et al. 2012; He et al. 2020; Scott et al. 2023). Furthermore, these HMR patterns are specifically enriched for genetic contribution to the heritability of cell-relevant clinical traits and diseases.

A number of epigenetic features have been used to identify islet-specific enhancers, including, but not limited to, enhancer-associated histone marks (e.g., H3K4me1 and H3K27ac), TF binding, and chromatin accessibility(Pasquali et al. 2014; Kycia et al. 2018; Varshney et al. 2017; Thurner et al. 2018; Greenwald et al. 2019; Cebola 2019; Chiou et al. 2021b; Rai et al. 2020; Kaplan et al. 2024). However, compared to more transient epigenetic states driven by dynamic changes in chromatin accessibility and histone modifications, our studies show that HMRs define both current and historically active enhancers. Therefore, HMRs hold the potential to provide a unique record of cell developmental history, exposure, and function. To date, methylation studies in islets have largely focused on comparing the methylomes of islets isolated from organ donors with and without T2D, highlighting distinct DNA methylation patterns associated with the disease. Notably, altered DNA methylation at cell-type specific regulatory elements was specifically associated with changes in gene expression related to β cell dysfunction and apoptosis, as well as TF binding, in islets from individuals with T2D versus controls. Despite these important findings, the majority of existing methylation studies have either profiled a smaller proportion of the islet genome or focused on differentially methylated regions (DMRs) linked to T2D(Yang et al. 2012, 2011; Ling et al. 2008; Rönn and Ling 2015; Avrahami and Kaestner 2019; Neiman et al. 2017; Dayeh et al. 2014; Thurner et al. 2018; Volkmar et al. 2012). Here, we sought to develop a comprehensive profile of the islet methylome that specifically captures genome-wide hypomethylation patterns in islets from individuals without diabetes.

In this study, we developed a workflow that integrates DNA hypomethylation with genotype-phenotype data to characterize the islet methylome and identify T2D-associated, islet-specific enhancer regions (**Fig. 1A**). We generated DNA methylation profiles from islets of donors without diabetes using whole-genome bisulfite sequencing (WGBS) and curated a reference dataset of islet HMRs. Using DNA methylation data from diverse cell types, we identified a subset of ∼4,800 unique islet HMRs and demonstrate their strong relationship to genes and TFs linked to β cell function. Further supporting their distinct function, islet-specific HMRs are highly enriched for known T2D risk variants, and partitioned heritability analysis also showed that islet HMRs are more enriched for genetic contribution to T2D heritability than HMRs from other functionally related tissues. We subsequently leveraged electronic health record (EHR)-linked genetic data from the Vanderbilt University Medical Center DNA biobank, BioVU, to evaluate the clinical and functional relevance of islet-specific HMRs. Using this resource, we developed an “HMR-WAS” for T2D, which focuses on variants in HMRs rather than across the entire genome. This approach allowed us to detect T2D-associated signals that would not have been detected above the genome-wide significance threshold in our smaller BioVU cohort of ∼17,000 subjects. Finally, we discovered three T2D-associated, islet-specific HMRs with enhancer activity that map to genes involved in insulin secretion and glucose metabolism: *PAM*, *SLC2A2*, and *GCK.* We demonstrate the regulatory capacity of these HMRs and the impact of T2D-associated variants on their activity using reporter assays in two rodent β cell lines. Ultimately, this integrated framework allowed us both to pinpoint islet enhancers of critical importance to islet cell identity, as well as to increase the visibility of functional variants linked to T2D.

**Figure 1.**
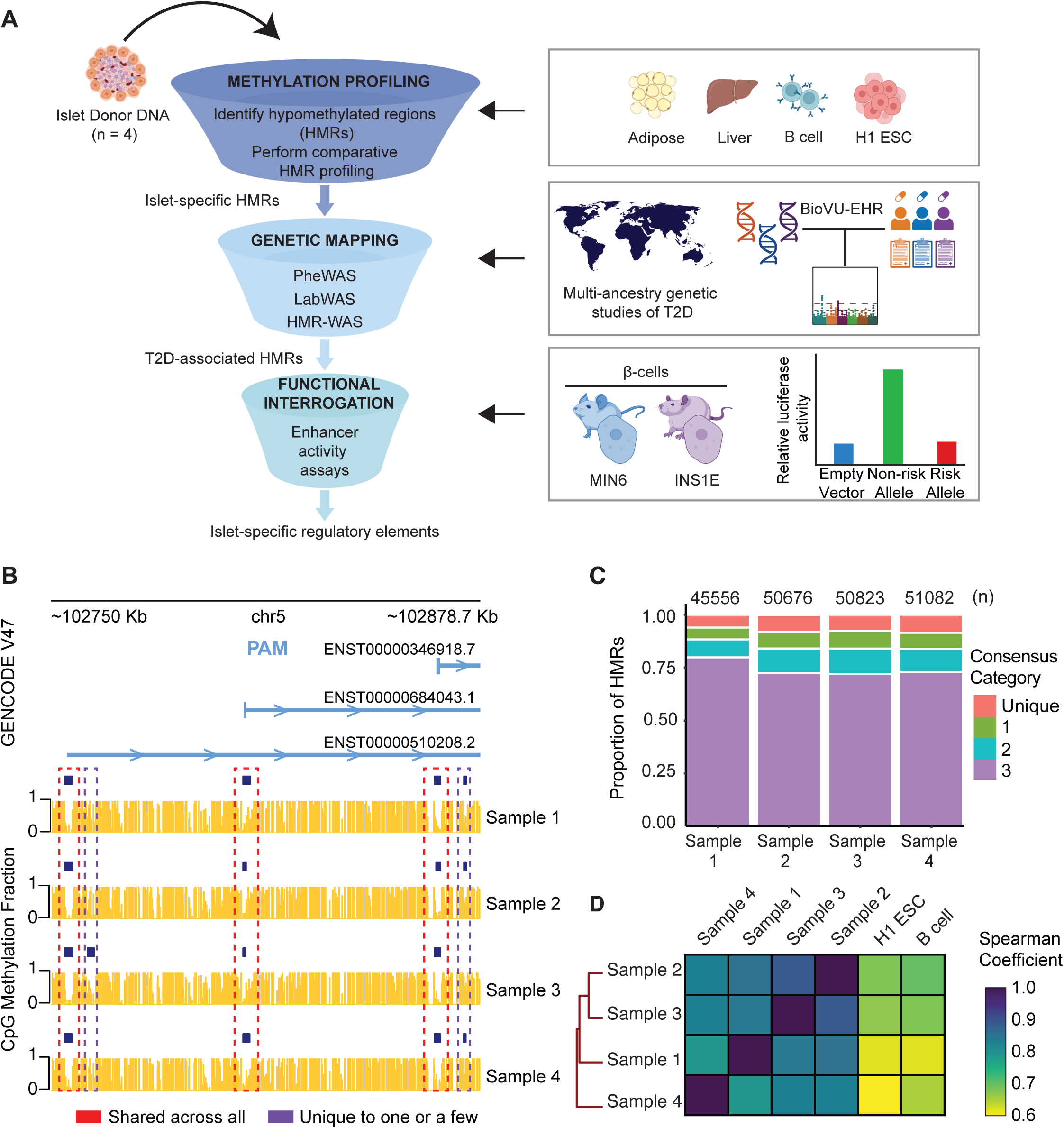
Islet WGBS data from non-T2D donors captures a consensus set of islet HMRs. **(A)** Overview of study workflow divided into three main components: methylation profiling, genetic mapping, and functional interrogation. **(B)** Multiple alignment of islet WGBS methylation and HMR tracks across four non-T2D donors at the *PAM* locus. Three representative isoforms of *PAM* from the GENCODE genes track (version 47) are shown to depict the overall diversity of isoforms at the gene locus. Methylation tracks are represented by gold vertical bars showing methylation value on a 0 to 1 scale per CpG site. Methylation fraction is determined by dividing the number of reads with a methyl-C by the total number of reads spanning the CpG site. HMRs represent clusters of adjacent, lowly methylated CpG sites detected using a 2-state hidden Markov model and are depicted by dark blue horizontal bars. **(C)** Bar graph showing the proportion of islet HMRs grouped into four categories based on the extent to which they are shared among donors. For each donor, a consensus category of ‘3’ indicates the fraction of islet HMRs that overlap HMRs found in all other donors. Islet HMRs assigned to categories ‘2’ and 1’ overlap regions present in two and one other donor(s), respectively; the category ‘Unique’ represents HMRs that are exclusive to a donor. **(D)** Heatmap depicting pairwise correlations across donors based on average methylation levels at reference islet HMRs. The color scale reflects the Spearman correlation value, and clusters were formed using the complete linkage method for hierarchical clustering. H1ESC and B-cell WGBS datasets are included as outgroups to highlight the strength of pairwise relationships between donors compared to cell types functionally unrelated to islets.

## RESULTS

### Whole-genome DNA methylation profiling reveals a consensus set of islet HMRs among non-T2D donors

To establish an islet reference methylome, we performed whole genome bisulfite sequencing (WGBS) on genomic DNA (gDNA) from pancreatic islets of four donors with no history of T2D (**Fig. 1B and Table S1**). A complete summary of sequencing information, alignment statistics, and coverage of symmetric CpG sites for all samples can be found in **Table S2**. We identified HMRs in each non-T2D donor using a 2-state hidden Markov model with beta-binomial distributions originally defined in Molaro et al. 2011 (Molaro et al. 2011; Song et al. 2020). We found that HMR features are highly similar across all donors with an average length of 1141 bp and 52 CpG sites (**Fig. S1A and S1B**). Likewise, the distribution of methylation levels at HMRs is also consistent, with a mean methylation value of 11.23% (**Fig. S1C**).

A comparison of donor HMR profiles revealed that over 70% of HMRs defined in a single individual overlap HMRs defined in all other individuals (**Fig. 1C and Table S3**). For each donor, there is little variability (*s* ≤ 1.05) in symmetric CpG coverage at HMRs by consensus categories, indicating that HMRs unique to one donor or shared with only one or two other individuals is not due to uneven or low sequencing coverage in our WGBS libraries (**Table S4**). HMRs identified in each donor that also overlap HMRs found in all other individuals were merged to create a consensus list of genomic regions totaling 35,721 HMRs, which we refer to henceforth as reference islet HMRs (**Fig. S1D-F**). Mean methylation values were determined for non-mutated symmetric CpG sites sequenced across all four donors (n = 29,954,671) to create a reference methylation and CpG coverage profile for islets (**Table S5**, **see Methods**).

Next, we measured the similarity in methylation levels across donors within this consensus HMR dataset and found that methylation levels are highly correlated across all pairwise comparisons (*ρ* ≥ 0.7, **Fig. 1D**). Furthermore, we observed weaker pairwise relationships (0.60 ≤ *ρ* ≤ 0.70) when applying methylation data from cell types functionally unrelated to islets (B cells and H1 ESC)(Scott et al. 2023). These results demonstrate that the high overall similarity between donors at reference islet HMRs is attributed specifically to methylation patterns measured at CpG sites in islets. When we compare methylation levels across donors within non-reference islet HMRs (n_HMR_ = 30,061), which includes HMRs that were not identified across all individuals, all pairwise correlation comparisons among donors reveal weak relationships (0.30 ≤ *ρ* ≤ 0.44), with even weaker ones for B cells and H1 ESC (**Fig. S1G and Table S3**). Taken together, we have determined a high-confidence reference islet HMR dataset, which excludes HMRs that were not defined across all donors, wherein this subset of regions and its methylation patterns are ‘representative’ of a non-T2D methylome.

### Identification of islet-specific HMRs reveals putative regulatory elements enriched for TF motifs and gene pathways regulating β cell identity and function

Previous work in our lab has shown that HMRs with varying levels of cell-type specificity can be defined by comparing the methylomes of diverse cell types and that these levels of specificity reflect unique developmental relationships and functions. In particular, this methodology can identify a subset of non-coding HMRs that are cell-type specific gene regulatory elements, including gene enhancers(Schlesinger et al. 2013; Scott et al. 2023; Hodges et al. 2011). Similarly, we compared HMR patterns between our reference islet methylome and other cell types and tissues representing various developmental timepoints and organ systems. These included publicly available WGBS data for adipose cells and liver for their interconnection to pancreatic islets in regulating whole-body glucose homeostasis, and unrelated cell types including H1 ESCs and B cells (**Table S6**). Of the four datasets included, liver, H1 ESC and B cell have all been analyzed in prior studies(Song et al. 2020, 2013; Molaro et al. 2011; Scott et al. 2023).

Hierarchical clustering of methylation levels from a merged HMR dataset comprised of HMRs defined across all cell types (n_HMR_ = 89,882) demonstrated that HMRs alone can recapitulate cell type relationships both distant and related to islets (**Fig. S2A**). This result is reinforced by the observation that the islet methylome forms a cluster with liver, reflecting the pancreatic islet-liver cross-talk based on reciprocal regulation of blood glucose levels through insulin and glucagon signaling(Röder et al. 2016). We also performed unsupervised *k*-means clustering to obtain groups of HMRs with similar methylation levels at merged HMRs, estimating the optimal number *of k*-means clusters as 6 (**Fig. S2B, see Methods**). We manually classified each *k*-means cluster based on represented cell types with average HMR methylation ≤ 50% for each cluster (**Fig. 2A**). The resulting heatmap revealed 6 different clusters of HMRs associated with cell- and lineage-specific gene regulatory activity based on stratification of top TF motifs enriched by *k*-means cluster (**Fig. S2C**). Notably, we identified an “Islet-specific” *k*-means HMR cluster, where islets alone have markedly lower methylation levels compared to the other cell types.

**Figure 2.**
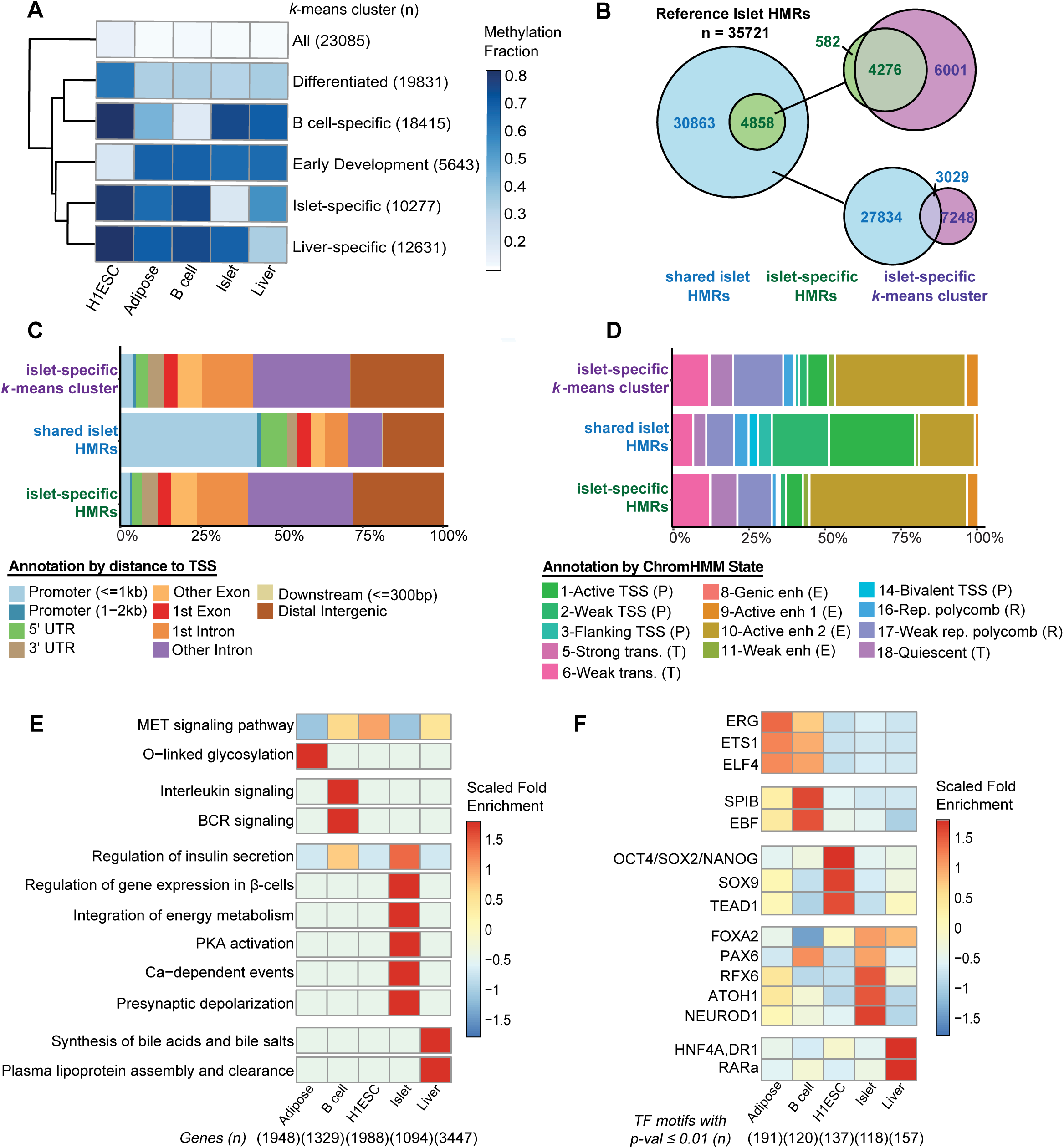
Comparative hypomethylation profiling identifies islet-specific HMRs enriched for TFs and pathways involved in islet function. **(A)** Heatmap showing *k*-means clustering of HMRs identified across cell types (n = 89,883) based on average CpG methylation levels per HMR. Each cluster was manually labeled to reflect biologically relevant cell type relationships. **(B)** Euler plot illustrating the overlap between cell-type-shared islet HMRs and islet-specific HMRs with HMRs represented in the “islet-specific” *k*-means group shown in panel A. Overlap was defined as a 1 bp minimum. **(C)** Bar graph of genomic feature annotations of cell-type-shared islet HMRs, islet-specific HMRs, and HMRs in the “islet-specific” *k*-means group. Annotations were assigned using the *ChIPseeker* R package. **(D)** Bar graph of HMR overlap with chromatin state annotations for the three HMR groups represented in panel B using a 13-state chromatin map for islets. Chromatin states are also labeled by their higher assigned category – enhancer (E), promoter (P) or transcribed (T)(Varshney et al. 2017). Overlap with ChromHMM annotations was determined using the *Bedtools intersect* software with default settings(Quinlan and Hall 2010). **(E)** Heatmap of top representative Reactome pathways for each cell-type specific HMR dataset. HMR-gene pairswere established using the nearestneighbor approach with the *ChIPseeker* R package(Yu et al. 2015). Scaled by row, color represents the ratio of the frequency of input genes with the associated term divided by the frequency of all genes annotated to that term. (**F**) Heatmap of top representative TFs for HMR groups represented in panel E. Scaled by row,color represents the fold change between the percentage of target regions with a target motif and the corresponding percentage in randomly selected background regions generated by the *HOMER* software package(Heinz et al. 2010).

The “islet-specific” *k*-means HMR cluster consists of 10,277 regions derived from a composite set of 89,882 HMRs merged across cell-types. As a result, some HMRs included within this *k*-means cluster may overlap HMRs called in multiple cell types. Because of this, we evaluated whether reference islet HMRs physically intersect HMRs defined in any of the other 4 cell types to identify a stringent set of islet-specific HMRs. This analysis enabled classification of reference islet HMRs into two categories: shared and islet-specific. Of the 35,721 reference-islet HMRs, 4,858 are islet-specific, or unique to islets, and the remaining 30,863 are shared with at least one other cell type (**Fig. 2B and Table S7**). Eighty-eight percent of islet-specific HMRs are captured within the “islet-specific” *k*-means HMR cluster shown in **Fig. 2A**, whereas only 10% of shared HMRs overlap this cluster (**Fig. 2B**). Both islet-specific HMRs and the “islet-specific” *k*-means HMR cluster are comprised of similar proportions of genomic features and functional annotations (**Fig. 2C and 2D**). Notably, ∼75% of islet-specific HMRs map to intronic or distal intergenic locations, as previously reported for cell-type specific HMRs (**Fig. 2C**)(Abascal et al. 2020; Panigrahi and O’Malley 2021; Schlesinger et al. 2013; Scott et al. 2023). In line with this finding, a 13-chromatin-state map for islets generated using ChromHMM for 33 cell types (including islets) demonstrated that ∼57% of islet-specific HMRs have chromatin states associated with enhancers(Varshney et al. 2017). In contrast, a majority of shared islet HMRs (43%) map to promoter locations, and 54% correspond to chromatin states associated with promoter activity (**Fig. 2C and 2D**). These results indicate a distinct functional role for islet-specific HMRs compared to those shared with other cell types.

Next, we used pathway enrichment analysis to compare the associated genes and biological pathways of islet-specific HMRs to the cell-type specific HMRs defined in adipose, B cell, H1 ESC and liver (**Fig. 2E**)(Croft et al. 2011). These results showed that islet-specific HMRs are highly enriched for nearest-neighbor genes that regulate gene expression in β cells. We also observe enrichment of processes controlling the multi-step insulin secretion pathway, including calcium-dependent events, PKA activation, and presynaptic depolarization(Rorsman and Ashcroft 2017). In line with our pathway enrichment results, islet-specific HMRs are enriched for the motifs of key islet TFs – FOXA2, RFX6, NEUROD1, and PAX6 – that are together responsible for endocrine cell differentiation, insulin gene transcription, and β cell maturation and function (**Fig. 2F**)(Lantz et al. 2004; Babu et al. 2008; Swisa et al. 2016; Naya et al. 1995; Walker et al. 2023; Smith et al. 2010; Ibrahim et al. 2024). In fact, 44-47% of islet-specific HMRs harbor the binding sites for these TFs. Furthermore, a recent study showed that *Rfx6* is a critical β cell hub gene required for stimulated insulin secretion, and its reduced expression in β cells was causally associated with early-onset T2D(Walker et al. 2023). Taken together, we have identified a subset of reference islet HMRs that are uniquely found in islets, the majority of which (≥ 50%) have enhancer chromatin states linked to associated genes essential for islet function and contain binding motifs for TFs that regulate β cell identity and function(Varshney et al. 2017). Consistent with these results, findings from both pathway and TF motif enrichment analyses show that islet-specific HMRs are depleted of biological processes and TF motifs associated with other cell-types.

### Islet-specific HMRs are highly enriched for SNPs linked to T2D and related traits

Integration of T2D GWA studies with functional genomic data have not only demonstrated that T2D risk variants are predominantly non-coding, but also enriched at islet enhancers, implicating perturbed islet transcription as a driving mechanism in T2D risk(Varshney et al. 2017; Cebola and Pasquali 2015; Pasquali et al. 2014; Walker et al. 2023; Gaulton et al. 2015; Parker et al. 2013; Scott et al. 2012). Accordingly, we asked whether islet-specific HMRs are enriched for GWAS loci associated with specific traits documented in the NHGRI-EBI GWAS catalog. Using a hypergeometric test, we determined that GWAS SNPs associated with traits “Type 2 Diabetes” (p-value = 2.30 × 10^−10^) and “Hemoglobin A1C Level*s*” (p-value = 6.50 × 10^−7^) are the most enriched in islet HMRs compared to other cell types and their cell relevant traits (**Fig. 3A, see Methods**)(Sollis et al. 2023; Bergman et al. 2020). We also discovered enrichment of traits that are not immediately attributable to islet function, including “Serum Creatinine Levels” (p-value = 2.62 × 10^−6^), “Hematocrit” (p-value = 1.07 × 10^−3^) and “Prostate Cancer” (p-value = 3.90 × 10^−5^)(Sollis et al. 2023). Multiple studies have shown each of these traits can serve as predictors to evaluate patients at higher risk for T2D given their close correlation to glucose uptake and insulin sensitivity in peripheral tissues(Bao et al. 2018; Song et al. 2022; Li et al. 2023; Harita et al. 2009; Kashima et al. 2017; Nayak et al. 2011; Qin et al. 2020; Merz and Thurmond 2020; Salazar-Vazquez et al. 2006; Tamariz et al. 2008; Nakanishi et al. 2004; Tulloch-Reid et al. 2004; Bansal et al. 2012; Pierce and Ahsan 2010; Kasper and Giovannucci 2006; Calton et al. 2007; Kasper et al. 2009). The enrichment of SNPs linked to T2D and clinical lab measures to diagnose and monitor T2D strongly implicates a functional role for variation within islet-specific HMRs in regulating genes critical to islet function and glucose homeostasis.

**Figure 3.**
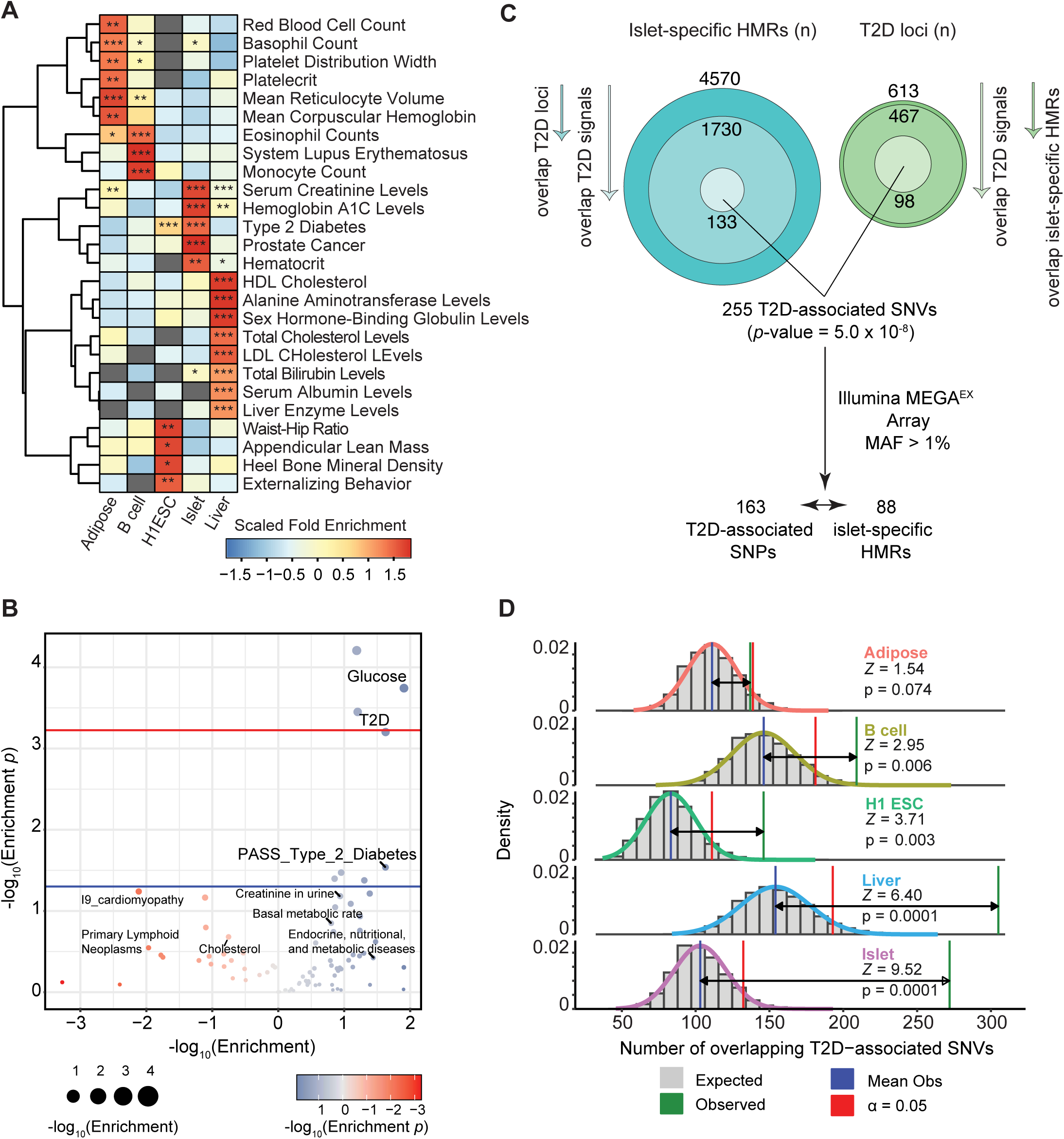
Islet-specific HMRs show a strong enrichment for variants associated with T2D and related endocrine traits. **(A)** Heatmap showing top traits from a trait enrichment analysis comparing SNP-trait associations in cell-type specific HMR datasets to those reported in the NHGRI-EBI GWAS catalog(Sollis et al. 2023). Scaled by row, color representsthefolddifferencebetween thefrequency of a trait within a cell-type specific HMR dataset divided by its frequency in the catalog. Significance was calculated using a hypergeometric test: *= *p*-value < 0.05, ** = *p*-value < 0.01, *** *p*-value < 0.001. Grey cells denote the absence of overlapping SNPs with an associated trait for a cell-type specific HMR group. **(B)** Volcano-style plot of S-LDSC partitioned heritability results across 84 traits for islet-specific HMRs. Thecolor and size of a point ^represent the -log^_10_*^p^*^−value of the^ for a trait and the corresponding ^(-)log^_10_^enrichment value, respectively^. For traits with an enrichment < 0, the output wasnegated following log-transformation and then plotted. A subset of points with *p*-value < 0.05 are labeled as well as those that are enriched (enrichment > 5) or depleted (enrichment < -5). The blue and red lines indicate the nominal (*p*-value = 0.05) and Bonferroni-corrected (*p*-value < 5.95 × 10^−4^) significance thresholds, respectively. **(C)** Euler plots showing the overlap between islet-specific HMRs and T2D loci identified collectively from the DIAMANTE and TDGGI studies as well as the genome-wide significant T2D signals at those loci(Mahajan et al. 2022; Suzuki et al. 2024). **(D)** Plot of the results of a permutation test evaluating the association between cell-type specific HMR datasets and genome-wide significant T2D association signals (n_T2D SNV_ = 83,690) measured in either or both DIAMANTE and T2DGGI studies(Mahajan et al. 2022; Suzuki et al. 2024). The grey histogram represents the expected distribution with a fitted normal curve in black. The blue, green, and red bars represent the mean overlap of the randomization evaluations, the observed overlap for the original HMR dataset, and a significant limit of 0.05, respectively.

We explore this further using GWAS summary statistics to perform HMR-stratified linkage disequilibrium score regression (S-LDSC) (**Table S8).** Using summary statistics from 84 traits and clinical lab values, we measured SNP-based heritability partitioned by islet-specific HMR annotations to establish the proportion of heritability concentrated within HMRs for each trait and lab(Finucane et al. 2015). We found SNP-based heritability of T2D-relevant traits to be significantly enriched within islet-specific HMRs (**Fig. 3B and S3**), reaffirming the impact of variation within islet-specific HMRs on islet dysfunction (Wysham and Shubrook 2020; Lv et al. 2022; Cohrs et al. 2020). These traits include “Glucose”, “T2D” and “PASS_Type_2_Diabetes”, with “Glucose” attaining the threshold for Bonferroni-corrected statistical significance and “T2D” at its borderline (p-value < 5.95 × 10^−4^, n_TRAIT_ = 84). Furthermore, “T2D” and “PASS_Type_2_Diabetes”, which are derived from separate GWA studies, are only nominally enriched or in some cases depleted in the non-islet specific HMRs (**Fig. S3**). While not significant by *p*-value (p-value < 0.05), “Creatinine in urine”, “Basal metabolic rate”, and “Endocrine, nutritional, and metabolic diseases” all achieve a positive enrichment value greater than 5. Similarly, SNP-based heritability of traits that do not involve islet function is depleted in islet HMRs (enrichment level < -5), including, but not limited to, “I9_Cardiomyopathy”, “I25_chronicIHD”, and “Menopause_Age”. These findings corroborate our expectation that genomic variation residing in islet-specific HMRs may specifically contribute to T2D heritability.

### Islet-specific HMRs significantly overlap T2D genetic loci informed by multi-ancestry T2D GWAS meta-analyses

Given these results, we subsequently leveraged summary statistics from partly independent multi-ancestry GWAS meta-analyses of T2D conducted by the DIAMANTE (Diabetes Meta-Analysis of Trans-Ethnic association studies) Consortium and T2DGGI (The Type 2 Diabetes Global Genomics Initiative) to further quantify the overlap between islet-specific HMRs and T2D loci (**Fig. 3C**)(Mahajan et al.; Suzuki et al. 2024). The combination of these two studies expanded the number of known T2D risk loci by more than 2.2-fold. Approximately 38% of islet-specific HMRs (1,730 out of 4,570) overlap with 613 T2D loci identified from both studies (see **Methods**). This in turn represents 76% of all T2D loci (467 out of 613) that contain at least one islet-specific HMR (**Fig. 3C and Table S10**). These loci are on average a mega-base and more in length due to linkage disequilibrium (LD) of the associated single nucleotide variants (SNVs) within the loci, compared to the more discrete and experimentally defined boundaries of HMRs. Therefore, we further filtered these loci to select only those with HMRs that specifically contain SNVs with rsID numbers and robust T2D association signals reaching genome-wide significance (p-value < 5.0 × 10^−8^), as measured in either or both studies. This filtering step revealed that ∼21% of the T2D loci that overlap islet-specific HMRs (98 out of 467) contain genome-wide significant T2D association signals. More specifically, these 98 loci encompass 133 islet-specific HMRs that collectively harbor 255 genome-wide significant T2D-associated SNVs (**Table S9**).

We next performed permutation tests to determine whether islet-specific HMRs overlap genome-wide significant T2D-associated SNVs identified in the DIAMANTE and T2DGGI studies (n_T2D SNV_ = 83,690) more often than expected for a size-matched set of randomly distributed genomic regions (**Fig. 3D**). The observed overlap was significantly greater than expected, exceeding the expected value of 103 by roughly 2.6-fold (p-value = 9.99 × 10^−5^). Notably, the overlap with T2D-associated SNVs is strongest in islets (*Z*_Islet_ = 9.52) compared to the permutation test results for the other cell type-specific HMRs. While liver-(p-value = 9.99 × 10^−5^, *Z*_Liver_ = 6.40), H1 ESC-(p-value = 2.99 × 10^−3^, *Z*_H1 ESC_ = 3.71) and B cell-specific HMRs (p-value = 5.69 × 10^−3^, *Z*_B cell_ = 2.95) are also significantly enriched for T2D-associated SNVs, the significance is orders of magnitude less than that of islet-specific HMRs. Furthermore, adipose-specific HMRs do not significantly overlap T2D-associated SNVs (p-value = 0.074, *Z*_Adipose_= 1.54). These findings for islet-, H1 ESC- and adipose-specific HMRs are corroborated by those obtained from the hypergeometric tests overlapping HMRs with SNP-trait associations in the NHGRI-EBI GWAS catalog (**Fig. 3A**). Conversely, liver- (FC (fold change) = 0.59, p-value = 0.97) and B cell- (FC = 1.16, p-value = 0.26) specific HMRs did not show significant enrichment of T2D GWAS SNPs in our trait enrichment analysis using this catalog. These results could be due to the T2D-associated signal count in the NHGRI EBI-GWAS catalog being ∼37-fold less than that of the DIAMANTE and T2DGGI studies combined. On the other hand, liver-specific HMRs are enriched for SNPs linked to “Hemoglobin A1C Levels”, which is a common clinical lab measure to diagnose and monitor T2D progression (FC = 2.32, p-value = 4.7 × 10^−3^). Collectively, we demonstrate that islet-specific HMRs form a distinct group of functional sequences that significantly overlap T2D- associated signals more frequently than other T2D disease-relevant tissues such as liver and adipose tissue. These results coincide with current evidence that show islet enhancers have a higher level of enrichment for T2D GWAS SNPs compared to enhancers of other tissues(Viñuela et al. 2020; Parker et al. 2013; Trynka et al. 2013).

### T2D-associated islet-specific HMRs show a strong association to metabolic phenotypes in phenome-wide scans

Mining EHR-linked patient genotype data provides a powerful opportunity to uncover previously unknown genetic associations between non-coding regions in the genome and clinical phenotypes and laboratory measures. We therefore performed phenome-wide (PheWAS) association scans for T2D-associated SNPs (MAF > 1%) overlapping islet-specific HMRs to better contextualize their clinical and functional roles (**Fig. 3C, see Methods**). Using another independent dataset filtered for genetic similarity to a European reference population (*N* = 66,278), we tested a total of 1,749 clinical outcomes against 163 T2D-associated SNPs that reside in islet-specific HMRs (**see Methods, Fig. 3C, and Table S11**)(Carroll et al. 2014; Denny et al. 2010, 2013). Overall, we found 22 phenotypes significantly associated with 19 SNPs across 12 islet-specific HMRs, totaling 59 statistically significant SNP-phenotype associations (2.82 × 10^−4^ ≤ p-value ≤ 1.25 × 10^−10^) **(Fig. S4A and Table S11**). About 58% (7 out of 12) harbor at least one SNP associated with a metabolic phenotype, and 83% (10 out of 12) exhibit enhancer-like chromatin features(Varshney et al. 2017). Associated nearest neighbor genes of these 12 HMRs ranked in descending order by the number of SNP-phenotype associations found at these loci are: *HLA-DRB5* (37), *HNF1B* (5), *PAM* (3), *ADCY5* (2), *AUTS2* (2), *CKB* (2)*, HIVEP2* (2)*, MAU2* (2), *BAG5* (1)*, HNF4A* (1), *PPIF* (1), and *RMST* (1) (n_GENE_ = 12).

Across the 17 phenotype categories in our data, the highest number of significant associations were observed in the endocrine/metabolic (28/59), dermatologic (12/59), and neoplasms (5/59) groups (**Fig. S4B**).

Not surprisingly, an islet-specific HMR within the highly polymorphic *MHC* locus -- also known as the Human Leukocyte Antigen (HLA) region, a major contributor to autoimmune diseases like type 1 diabetes (T1D) and rheumatoid arthritis -- has the highest number of SNP-phenotype associations (n_SIGNAL_ = 37) (**Fig. S4C**). These signals span four SNPS in complete LD (r^2^ = 1.0) in the European subpopulation from Phase 3 of the 1000 Genomes Project. Residing in the 5’UTR of the class II MHC gene *HLA-DRB5* (major histocompatibility complex, class II, DR beta 5), this HMR’s SNPs are confirmed eQTLs for *HLA-DRB5* in multiple tissues that primarily govern blood sugar and whole-body energy metabolism, including subcutaneous adipose tissue, skeletal muscle, and thyroid(Lonsdale et al. 2013). Additionally, the gene regulatory influence of this region is supported by its assigned chromatin states associated with transcriptional regulation(Varshney et al. 2017). Within the endocrine/metabolic group, all SNPs were significantly associated with T2D-related phenotypes, including “Insulin pump user” (7.09 × 10^−7^ ≤ p-value ≤ 8.95 × 10^−7^) and “Diabetes mellitus” (1.42 × 10^− 7^ ≤ p-value ≤ 1.73 × 10^−7^). Other associated traits include autoimmune diseases “Type 1 diabetes” (1.54 × 10^−10^ ≤ p-value < 2.71 × 10^−10^), “Multiple sclerosis” (1.25 × 10^−10^ ≤ p-value ≤ 2.27 × 10^−10^), and “Psoriasis vulgaris” (2.22 × 10^−5^ ≤ p-value ≤ 2.37 × 10^−5^).

Beyond the *MHC* locus, we found 14 phenotypes significantly associated with 15 SNPs across 11 islet-specific HMRs, totaling 22 statistically significant SNP-phenotype association signals (6.06 × 10^− 5^ ≤ p-value ≤ 2.48 × 10^−7^) (**Fig. S4D**). The highest proportion of hits corresponded to the endocrine/metabolic (7/22), neoplasms (5/22), and mental disorders phenotype groups (3/22). Approximately 82% of the 11 islet-specific HMRs (9 out of 11) located outside of the *MHC* locus have enhancer-like chromatin features(Varshney et al. 2017). Among this subset of nine HMRs are regions that map to genes *PAM* (peptidylglycine alpha-amidating monooxygenase), *CKB (*creatine kinase B), and *HNF4A* (hepatocyte nuclear factor 4). Non-synonymous coding variants in *PAM* have been previously implicated in both increased T2D risk and reduced insulinogenic index(Huyghe et al. 2013; Steinthorsdottir et al. 2014; Lek et al. 2016; Thomsen et al. 2018). The non-coding HMR near the *PAM* locus harbors three metabolic-related association signals linked to the rs76177300-SNP: “Nonsenile Cataract” (p-value = 5.39 × 10^−5^, OR = 3.97), “Diabetes mellitus” (p-value = 5.71 x -10^−5^, OR = 1.17) and “Type 2 diabetes” (p-value = 6.06 x -10^−5^, OR = 1.17). These results align with genetic studies showing that shared genetic overlap between T2D and cataracts is driven by T2D-specific variants(Jiang et al. 2024; Li et al. 2014). The rs11626787-SNP, located in a distal intergenic HMR near the *CKB* locus, is associated with decreased risk for circulatory system trait “Other hypertensive complications” (p-value = 4.67 × 10^−5^, OR = 0.85) and mental disorders trait “Psychogenic and somatoform disorders” (p-value = 5.21 × 10^−5^, OR = 0.74). Given *CKB* is involved in thermogenic respiration, a process that helps counteract obesity and metabolic disease, the *CKB*-affiliated HMR may exert protective effects on conditions beyond the endocrine system(Rahbani et al. 2021; Carpentier et al. 2018). Finally, HNF4A, a key TF primarily influencing insulin production in islets, harbors the rs6103716-SNP located within an intronic HMR(Ng et al. 2024; Maestro et al. 2007). This SNP is linked to increased risk for “Type 2 diabetes with neurological manifestations” (p-value = 2.52 × 10^−5^, OR = 1.15). Taken together, our phenome-wide scans demonstrated that most phenotype association signals for a subset of T2D-associated islet-specific HMRs (n_HMR_ = 12, n_T2D SNP_ = 19) are metabolic/endocrine-related, including, but not limited to, “Type-1 diabetes”, “Insulin pump user” and “Hypothyroidism”. These results also highlighted the potential pleiotropic effects of these HMRs, whereby their gene regulatory effects are linked to biological outcomes underlying risk of clinical phenotypes beyond T2D and other metabolic-related traits.

### T2D-associated islet-specific HMRs show a strong association to metabolic clinical laboratory tests in lab-wide scans

LabWAS models genetic associations to clinical lab measures instead of clinical phenotypes (i.e., IC9/ICD10 diagnostic codes mapped to phecodes). EHR-based quantitative lab tests, such as glucose and HbA1C levels in the blood, are not only important clinical markers of disease, but are also direct biochemical measurements of underlying cell biology. Thus, we leveraged clinical lab data from the same BioVU cohort above (see previous section) to explore the relationship between genetic variants at T2D-associated islet-specific HMRs and laboratory-derived data (**Fig. 3C, Table S12 and see Methods**)(Goldstein et al. 2020; Dennis et al. 2020). For the 308 laboratory traits tested, we found 17 labs significantly associated with 43 SNPs across 22 islet-specific HMRs, totaling 62 statistically significant SNP-lab association signals in all (5.79 × 10^−5^ ≤ p-value ≤ 5.52 × 10^−59^) (**Fig. S5A and Table S12**). About 45% of these HMRs (10 out of 22) harbor at least one SNP with a genetic association to a metabolic clinic lab measure, and 95% have enhancer-like chromatin features (21 out of 22)(Varshney et al. 2017). Associated nearest neighbor genes of these HMRs ranked in descending order by the number of SNP-lab trait associations found at these loci are: *GCK* (10), *HLA-DRB5* (9), *LOCI100130744* (5), *MAST2* (4), *RMST* (4), *TMCC2* (4), *ZBTB38* (4), *CKB* (3), *RASGRP1* (3), *ADCY5* (2), *ANKDD1B* (2), *FOXN3-AS2* (2), *GLIS3* (2), *KLHL42* (2), *MAU2 (*2), *BAG5* (1), *LINC00575* (1), *SLC2A2* (1), and *XRCC3* (1) (n_GENE_ = 19). Across the 12 categories of labs, we found that most laboratory-trait association signals were linked to a metabolic clinical-lab test (35/62), followed by blood-cell traits (19/62), cardiovascular-(5/62) and immune-related (3/62) tests (**Fig. S5B**). The most significant associations across these laboratory-test groups were: “Glucose lab (Gluc)” (rs1799884-T, p-value = 3.88 × 10^−25^, OR = 1.07), “Platelet mean volume in blood (MPV)” (rs4400674-A, p-value = 5.52 × 10^−59^, OR = 0.90), “Creatine kinase–MB/Total creatine kinase in Serum or Plasma (MBRat)” (rs7693-T, p-value = 8.06 × 10^−10^, OR = 1.08), and “Leukocytes [#/volume] in Blood by Automated count (WBC)” (rs2523506-T, p-value = 7.22 × 10^−7^, OR = 1.03). The four SNPs listed here reside in the *GCK*, *TMCC2*, *BAG5*, and *HLA-DRB5* loci, respectively.

*HLA-DRB5* and *GCK* (glucokinase), both of which harbor pathogenic variants (loss of function and missense) that are associated with elevated T2D risk, are the top two genes based on the number of laboratory-trait association signals found at these loci (*HLA-DRB5*: n_SIGNAL_ = 10; *GCK*: n_SIGNAL_= 9)(Zhao et al. 2017; Schiabor Barrett et al. 2024). In fact, the same four SNPs with phenotype associations that map to an islet-specific HMR found at the 5’UTR of *HLA-DRB5* (see previous section) all harbor genetic links to the blood-cell lab trait “Erythrocyte distribution width [Ratio] by Automated count” (p-value ≤ 2.30 × 10^−5^) and metabolic lab measure, “Protein serum/plasma” (p-value ≤ 6.55 × 10^−6^). For *GCK*, both “Glucose (Gluc)” and “Hemoglobin A1C (Glycated)” lab measures are significantly associated with multiple SNPs: two located in intron 1 (rs2908289 and rs730487; 4.00 × 10^−5^ ≤ p-value ≤ 7.87 × 10^−25^) and three in the promoter region (rs2268570, rs2971670, and rs1799884; 3.88 × 10^−25^ ≤ p-value ≤ 3.27 × 10^−5^). Additionally, the chromatin state associated with the entire islet-specific HMR found at intron 1 of *GCK* is “10_Active_enhancer_2”, while the islet-specific HMR located at the promoter proximal site (≤ 1kb TSS) has both enhancer- and promoter-specific chromatin features(Varshney et al. 2017). The genetic associations to metabolic-related labs within the *HLA-DRB5*- and *GCK*-affiliated islet-specific HMRs point to a regulatory role for these regions in pathways underlying islet dysfunction and T2D.

We also observed other loci, in addition to *MHC*, that harbor both phenotype- and laboratory-trait association signals at T2D-associated islet-specific HMRs. In fact, *CKB*, *MAU2* (MAU2 sister chromatid cohesion factor), and *ADCY5* (adenylate cyclase 5) meet these criteria, and the HMRs residing within these loci exhibit chromatin states indicative of active enhancers (i.e., “10_Active_enhancer_2”)(Varshney et al. 2017). Beyond its genetic links to mental and circulatory system disorders identified in our phenome-wide scans, the rs11626787-SNP within the *CKB*- affiliated HMR is also associated with increased risk of cardiovascular lab measure, “Creatine kinase–MB/Total creatine kinase in Serum or Plasma (MBRat)” (p-value = 5.85 × 10^−8^, OR = 1.07). For *ADCY5*, the rs11708067-SNP found within the 5’UTR is not only associated with both decreased risk of “Diabetes mellitus” (p-value = 4.39 × 10^−7^, OR = 0.90) and “Type 2 diabetes” (p-value = 2.48 × 10^−7^, OR = 0.90), but also whole blood glucose (p-value = 6.49 × 10^−13^, OR = 0.96). Finally, the *MAU2*- affiliated exonic HMR harbors a SNP that is associated with both increased risk of “Diabetes mellitus” (rs11085259-T: p-value = 6.62 × 10^−6^, OR = 1.10) and “Type 2 diabetes” (rs11085259-T: p-value = 1.06 × 10^−6^, OR = 1.11). Interestingly, though, our LabWAS results showed that this same SNP could have a protective effect on metabolic-lab measures “Triglyceride [Mass/volume]” (p-value = 6.76 × 10^−6^, OR = 0.96) and “Cholesterol [Mass/volume] in serum or plasma” (p-value = 1.26 × 10^−6^, OR = 0.95). Taken together, the findings from our phenome- (PheWAS) and lab-wide (LabWAS) scans in the BioVU EU cohort support a clinical and functional role for a subset of T2D-associated islet-specific HMRs, where genetic variation disrupting their regulatory activity contributes to T2D and related metabolic traits.

### HMR-WAS reveals T2D risk variants impacting putative enhancer of *PAM*

We show that T2D signals from two of the largest T2D GWA studies map disproportionately to islet-specific HMRs (**Fig. 3D**). Importantly, these studies pool multiple T2D GWAS datasets to increase the size of their T2D case-control cohort, thereby improving statistical power to detect novel T2D association signals(Mahajan et al.; Suzuki et al. 2024). This is especially important when considering the variants with modest associations to T2D that would be undetected in smaller cohorts. Given our results, we hypothesized that restricting the analyzed SNPs to those falling only within HMRs would boost our statistical power, allowing us to detect biologically meaningful signals obscured in a typical GWAS. To test this idea, we developed a deviation of a GWAS, called “HMR-WAS”, a region-based approach that limits association tests to variants found in islet-specific HMRs, rather than genome-wide markers (**Fig. 4A**).

**Figure 4.**
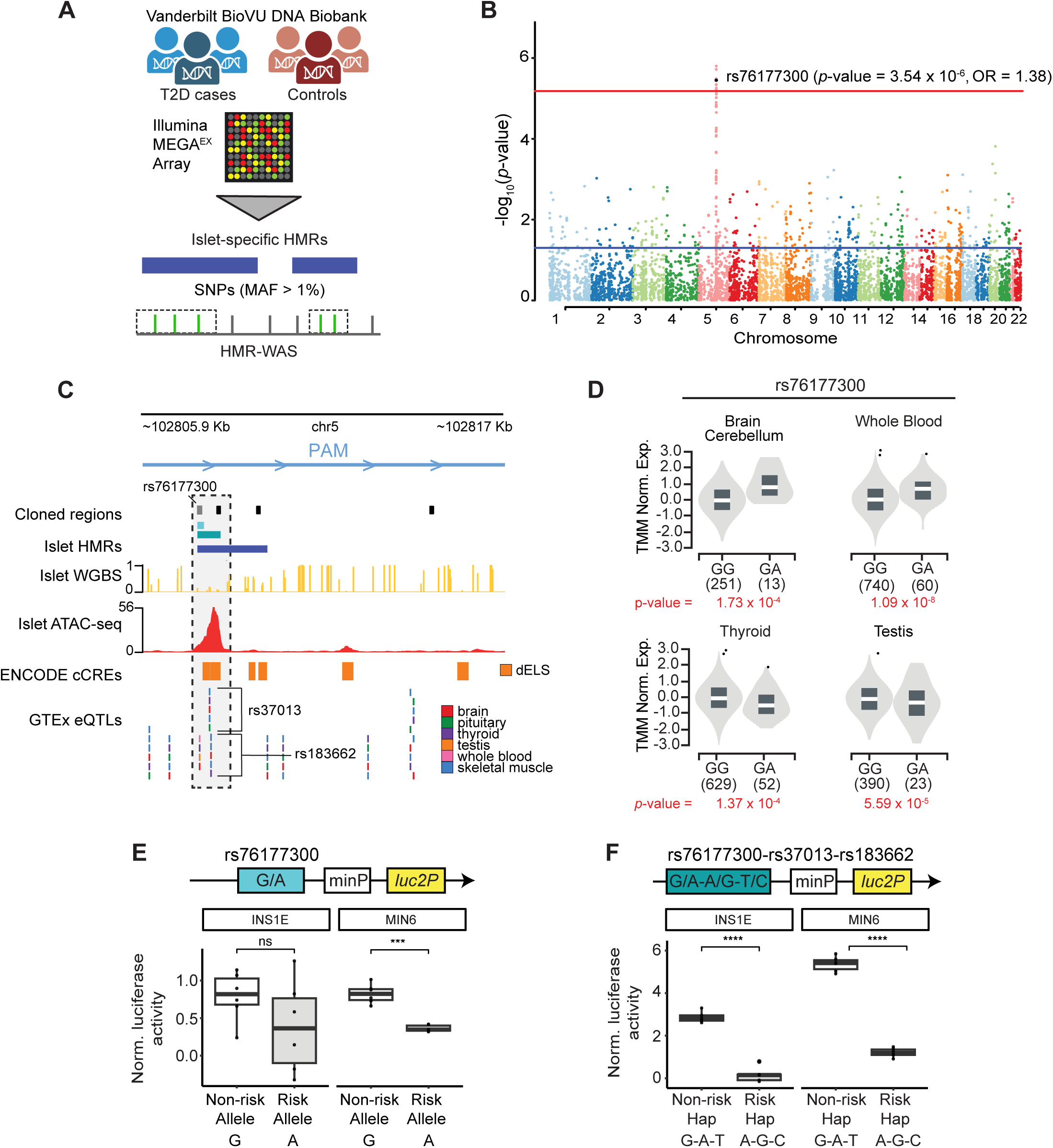
HMR-WAS reveals a T2D signal at an islet-specific HMR with enhancer activity surrounding *PAM* locus. **(A)** Overview of the cohort and pre-processing steps of an HMR-WAS for T2D in BioVU. Markers on the Illumina MEGA^EX^ array that overlap islet-specific HMRs and have a MAF > 1% were prioritized for association testing with the T2D phecode (250.2). **(B)** Manhattan plot of HMR-WAS results for T2D. The x-axis represents chromosomal position, and the y-axis shows the -log_10_(*p*-value) for each SNP-T2D association. The red and blue lines indicate the Bonferroni threshold (*p*-value < 6.86 × 10^−6^) and a *p*-value of 0.05, respectively. The black dot denotes the representative SNP within an LD peak. **(C)** Functional annotation of the *PAM*-associated islet-specific HMR harboring the SNP highlighted in panel B. The islet WGBS track represents the average methylation levels at non-mutated symmetric CpG site across islet donors, and the islet ATAC-seq data corresponds to sample ‘Islet 1’ in Varshney et al. 2017. **(D)** Violin plots showing normalized *PAM* expression associated with the rs76177300-eQTL in several GTEx tissues(Lonsdale et al. 2013). The x-axis displays genotype groups with corresponding sample counts. **(E)** Box plot illustrating functional validation of 200-bp segments flanking rs76177300. The risk allele (A) exhibited decreased transcriptional activity compared to the non-risk allele (G) in INS1E and MIN6 β-cells. Results on the y-axis are presented as the fold-change in Firefly:*Renilla* luciferase activity relative to the empty vector control, analyzed via unpaired two-sided *t*-tests (n = 6 per allele). **(F)** Box plot illustrating functional validation of 711-bp haplotype segments comprising rs76177300 and nearby *PAM* GTEx eQTLs. The T2D risk haplotype (A-G-C) exhibited lower transcriptional activity than the non-risk haplotype (G-A-T) in both cells. Haplotypes were defined using European population data from 1000 Genomes Project (Phase 3). Calculation methods are identical to those in panel E (n = 6 per haplotype); * = *p*-value < 0.05; ** = *p*-value < 0.01; *** = *p*-value < 0.001.

We performed both a standard GWAS and HMR-WAS for T2D using a study cohort from BioVU, which included 4,656 T2D cases and 12,748 control subjects of European descent (*N* = 17,404). All T2D cases and controls were identified using the Northwestern University Type 2 diabetes mellitus (T2DM) algorithms, which applies stringent criteria to differentiate between T2D cases and controls based on different types of EHR data (e.g., diagnoses, medications, and lab test results)(Kho et al. 2012; Wei et al. 2012; Hripcsak et al. 2019; Pacheco and Thompson). For our T2D HMR-WAS, we further filtered for variants that have a minor allele frequency (MAF) greater than 1%, resulting in 7,290 SNPs (all autosomal) considered for association testing to T2D (**Table S13**). Our HMR-WAS results revealed a region-wide significant T2D risk signal associated with the rs76177300-SNP (rs76177300-A: odds ratio = 1.38; p-value = 3.54 × 10^−6^; MAF = 4.573%, Bonferroni-corrected p-value < 6.86 × 10^−5^) that did not reach statistical significance via false discovery rate (FDR) or Bonferroni correction in our standard GWAS (**Fig. 4B, S6A-6C, and Tables 13-14**). By adding SNPs in LD with rs76177300 within a 1000-kb window (r^2^ ≥ 0.1), we achieved an LD peak indicating a region of strong association surrounding the rs76177300-SNP in our T2D HMR-WAS. In contrast, our standard T2D GWAS identified a large LD block of 64 SNPs in high LD (r^2^ ≥ 0.817, European subpopulation from Phase 3 of the 1000 Genomes Project) that spans ∼33.9-kb and maps to *TCF7L2* (transcription factor 7-like 2), the most potent and well-documented genetic locus linked to T2D to date(Del Bosque-Plata et al. 2021; Sladek et al. 2007; Grant et al. 2006). Of note, this signal is absent in our HMR-WAS because the associated SNPs do not overlap islet-specific HMRs.

The HMR-WAS rs76177300-SNP was previously associated with T2D (odds ratio = 1.10; p-value = 1.0 × 10^−10^) in a GWAS using genetic data from >400,000 White British participants of the UK Biobank (18,945 T2D cases and 388,756 controls)(Gagliano Taliun et al. 2020; Bycroft et al. 2018). Additionally, this variant is corroborated by genome-wide significant T2D association signals detected in both T2D GWAS meta-analyses conducted by the T2DGGI and DIAMANTE Consortium (DIAMANTE: MR-MEGA p-value = 2.04 × 10^−24^, fixed-effects OR = 1.16; T2DGGI: MR-MEGA p-value = 2.94 × 10^−37^, fixed-effects OR = 1.11; **Table S9**)(Mahajan et al.; Suzuki et al. 2024). These results demonstrate that consideration of HMR context for prioritizing SNPs is effective in identifying true disease association signals that do not meet stringent genome-wide significance thresholds.

The rs76177300-SNP is located in an HMR upstream of *PAM*. Given the role of T2D-associated *PAM* risk alleles and *PAM* deficiency in β cell dysfunction, we postulated that the *PAM*-associated islet-specific HMR is a putative enhancer regulating its gene expression(Huyghe et al. 2013; Steinthorsdottir et al. 2014; Lek et al. 2016; Thomsen et al. 2018; Merkler et al. 2022). To investigate the regulatory potential of this region, we integrated our reference islet methylation profile with other functional genomics data (**Fig. 4C**). This included genomic tracks of islet ATAC-seq data, candidate *cis*-Regulatory elements (cCREs) derived from ENCODE data, and expression quantitative trait loci (eQTLs) identified from the Genotype-Tissue Expression (GTEx) Project(Varshney et al. 2017; Lonsdale et al. 2013; Abascal et al. 2020). Not only does an ATAC-seq peak overlap the islet-specific HMR at the *PAM* locus, but approximately 47% of the ∼2.1-kb region is spanned by four cCREs that all have distal-enhancer like signatures (dELS)(Abascal et al. 2020). ChromHMM annotations generated for human pancreatic islets corroborate the putative regulatory activity of the HMR (**Fig. S7A**). Furthermore, the rs76177300-SNP specifically localizes to the part of the islet-specific HMR labeled as “10_Active_enhancer_2”, which is associated with H3K27ac and H3K4me1 histone modifications and spans approximately 25.8% of the region(Varshney et al. 2017; Heintzman et al. 2009; Kang et al. 2021; Creyghton et al. 2010; spicuglia and Vanhille 2012; Rada-Iglesias et al. 2010). We also speculate that this region is linked to both enhancer and promoter activity given its genomic location relative to different *PAM* transcriptional isoforms (**Fig. 1B**). The functional relationship between this islet-specific HMR and *PAM* mRNA expression is supported by the presence of eQTL SNPs within the region, including rs76177300-SNP as an eQTL associated with *PAM* expression in brain (cerebellum), thyroid, whole blood, and testis (**Fig. 4C and 4D**). While not in LD with the rs76177300-SNP, two neighboring SNP eQTLs in high LD with each other (r^2^ = 0.829, European subpopulation from Phase 3 of the 1000 Genomes Project) - rs37013 and rs183662 - also associate with *PAM* expression in a variety of other tissues, including but not limited to, skeletal muscle, brain, pituitary and thyroid(Lonsdale et al. 2013).

To validate the regulatory potential of this islet-specific HMR and to understand the impact of genetic variation on its regulatory activity, we performed luciferase reporter assays in two widely used cellular models of the islet β cell: INS1E rat insulinoma and MIN6 mouse insulinoma cells. We cloned two different copies of the 200-bp DNA sequence surrounding the rs76177300-SNP: one containing the T2D risk allele (A) and the other, the non-risk allele (G) (**Fig. 4E and Table S15**). In MIN6 cells, the G non-risk allele exhibited significantly increased transcriptional activity compared to the A risk allele by 2.36-fold (**Fig. 4E**, p-value = 1.37 × 10^−4^). Equivalent data for INS1E cells was directionally consistent but not significant (p-value = 0.192). These data suggest that not only does this smaller portion of the HMR exhibit enhancer activity but that rs76177300-SNP acts as a functional variant affecting activity levels demonstrated in both islet cell lines.

The islet HMR encompasses several cCREs beyond the sequence immediately surrounding rs76177300 (**Fig. 4C**). This additional sequence likely contains regulatory DNA, including potential TF-binding motifs that contribute to enhancer activity. Furthermore, the region harbors two other *PAM*-associated eQTLs (rs37013 and rs183662) that could confer allele-specific effects on regulatory activity. Using Phase 3 haplotype data from the 1000 Genomes European population, we designed two different copies of the 711-bp DNA sequence surrounding rs76177300 and these neighboring SNPs: one with the T2D-associated risk haplotype containing the alternate alleles for each of the three SNPs (A-G-C) and the other the non-risk haplotype containing the reference alleles (G-A-T)(**Fig. 4F and Table S15**)(Auton et al. 2015; Machiela and Chanock 2015). In both cell lines, the AGC-risk haplotype had significantly decreased transcriptional activity compared to the GAT-non-risk haplotype (GAT-non-risk: INS1E, 18.44-fold, p-value = 2.35 × 10^−6^; MIN6, 4.54-fold, p-value = 5.09 × 10^−9^). A sequence spanning the transcription start site (TSS) of the *PAM* promoter region (-249, +1) served as a technical positive control and exhibited a mean 3.22- and 18.8-fold increase in transcriptional activity compared with an empty-vector in INS1E and MIN6 cells, respectively (INS1E: p-value = 1.47 × 10^−4^; MIN6: p-value = 1.90 × 10^−6^) (**Fig. S7B**). These results suggest that one or more of these variants cause decreased regulatory activity in INS1E and MIN6 cells. Altogether, our findings with both 200- and 711-bp stretches of DNA surrounding the rs76177300-SNP validate the enhancer activity of this islet-specific HMR and strongly implicate its regulation of *PAM* expression in islets.

### LabWAS reveals islet-specific, enhancer HMRs regulating glucose metabolism genes

We identified two islet-specific HMRs in proximity to genes that have essential roles in regulating blood glucose levels and are expressed in islet β cells: *GCK* and *SLC2A2* (glucose transporter solute carrier family member 2), which encodes GLUT2(Thorens et al. 1990; Abu Aqel et al. 2024). Together, both GCK and GLUT2 are integral proteins in a metabolic cascade of events that ultimately leads to the synthesis and secretion of insulin. Moreover, genetic variation in both genes have been previously implicated in T2D(Barroso et al. 2003; Fu et al. 2013; Muller et al. 2014; Alcolado et al. 1991; Laukkanen et al. 2005; YUN et al. 2019).

For *GCK*, we identified two SNPs found in intron 1 associated with increased whole blood glucose and glycated hemoglobin (**Fig. 5A**, rs2908289_Gluc: p-value = 7.87 × 10^−25^, OR = 1.07; rs2908289_ HgbA1C: p-value = 3.91 × 10^−5^, OR = 1.05; rs730497_Gluc: p-value = 1.71 × 10^−24^, OR = 1.07, rs730497_HgbA1C: p-value = 4.00 × 10^−5^, OR = 1.05). Integration of islet methylation and other functional genomics datasets at the *GCK* locus highlights an intronic, islet-specific HMR with putative regulatory activity based on overlapping islet accessible chromatin peaks and enhancer-like signatures assigned by ENCODE cCRE data and a 13-state ChromHMM model for pancreatic islets (**Fig. 5B and S7C**)(Varshney et al. 2017; Abascal et al. 2020). In addition, both rs2908289 and rs730497 SNPs are single-tissue eQTLs affecting *GCK* expression levels in thyroid (**Fig. 5B and 5C**).

**Figure 5.**
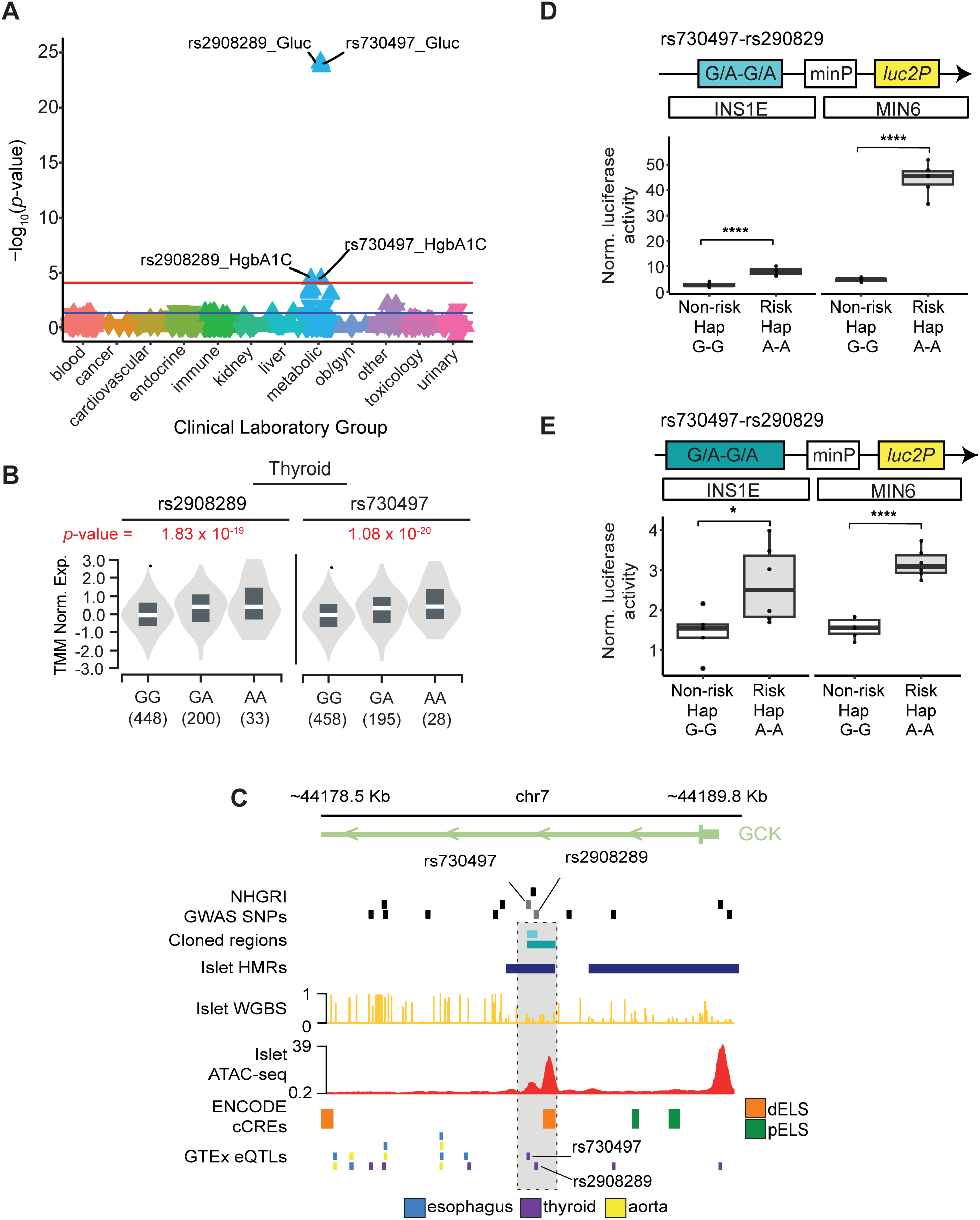
Islet-specific HMR with enhancer activity at *GCK* locus contains T2D-associated SNPs linked to endocrine labs. **(A)** Manhattan plot of laboratory-trait associations for T2D-associated SNPs rs2908289 and rs730497. The combined laboratory-trait associations for both SNPs are grouped and color-coded by clinical laboratory-test group on the x-axis and plotted by -log_10_(*p*-value) on the y-axis. Triangles point upward or downward to denote increased or decreased lab values with each minor allele copy. The red and blue lines indicate the Bonferroni-corrected threshold (*p*-value < 8.12 × 10^−5^) and a *p*-value of 0.05, respectively. (**B)** Multiple alignment of functional genomic data at the *GCK*-associated islet-specific HMR containing the SNPs shown in panel A. The islet WGBS track displays mean methylation levels at non-mutated symmetric CpG sites across islet donors, and the islet ATAC-seq data is derived from the “Islet 1” sample reported in Varshney et al. 2017. **(C)** Violin plots showing normalized *GCK* expression associated with the rs2908289- and rs730497-eQTLs in thyroid based on GTEx data(Lonsdale et al. 2013). The x-axis represents genotype groups and associated sample size. **(D)** Box plot representing functional validation of 286-bp haplotype segments containing rs2908289 and rs730497. The risk haplotype (A-A) exhibited significantly increased transcriptional activity compared to the non-risk haplotype (G-G) in INS1E and MIN6 β-cells. Results are plotted as a fold-change of Firefly:*Renilla* luciferase activity relative to the empty vector control. Statistical significance was calculated using an unpaired two-sided *t*-test (n = 6 per haplotype). **(E)** Box plot illustrating functional validation of 790-bp haplotype segments encompassing rs2908289, rs730497, and an ENCODE cCRE with a dELS. In parallel to our findings shown in panel D, the risk haplotype (A-A) showed greater transcriptional activity than the non-risk haplotype (G-G) in both cell lines. Relative luciferase activity and *p*-values were calculated using the same methods described in panel D (n = 6 per haplotype); * = *p*-value < 0.05; ** = *p*-value < 0.01; *** = *p*-value < 0.001.

To functionally examine this locus and the regulatory effects of residing T2D-associated SNPs, we performed luciferase reporter assays testing two different copies of the 286-bp DNA sequence surrounding the rs2908289-rs730497 SNPs: one containing the T2D risk haplotype (A-A) and the other the non-risk haplotype (G-G) (**Fig. 5D, Fig. S7D, and Table S15**). In both islet cell lines, both the A-A risk and G-G non-risk haplotype sequences exhibited increased transcriptional activity relative to the empty-vector. Furthermore, altering alleles of the rs2908289-rs730497 genotype resulted in differential allelic enhancer activity. More specifically, the A-A risk haplotype exhibited significantly increased transcriptional activity compared to the G-G non-risk haplotype by 3.2- and 9.3-fold in INS1E and MIN6 cells, respectively (INS1E: p-value = 4.90 × 10^−4^; MIN6: p-value = 1.30 × 10^−5^).

We also tested a larger stretch of DNA sequence within the HMR that encompasses a cCRE with a predicted distal enhancer-like signature (**Fig. 5E and Table S15**)(Abascal et al. 2020). In parallel to the results obtained from the shorter DNA sequence, the A-A risk haplotype showed significantly increased transcriptional activity compared to the G-G non-risk haplotype by 1.6- and 2.0-fold in INS1E and MIN6 cells, respectively (INS1E: p-value = 3.20 × 10^−4^; MIN6: p-value = 1.0 × 10^−4^). This result is supported by the same direction of effect on *GCK* gene expression for both rs2908289 and rs730497 SNP eQTLs, whereby normalized expression is significantly higher with the alternative allele or risk haplotype (A-A) compared to the reference allele or non-risk haplotype (G-G) (rs2908289: p-value = 1.83 × 10^−19^, NES = 0.37; rs730497: p-value = 1.08 × 10^−20^, NES = 0.39). Altogether, these results highlight the regulatory capacity of this intronic islet-specific HMR in the *GCK* gene as well the allele-specific enhancer activity of the rs2908289-rs730497 SNPs.

For *SLC2A2*, we identified an islet-specific HMR immediately upstream of the gene harboring an association signal for whole blood glucose (rs1905505_Gluc: p-value = 8.36 × 10^−10^; OR = 0.97). Although not statistically significant after multiple testing correction, the next strongest association signal is related to glycated hemoglobin (HbA1C) levels (HgbA1C: p-value = 1.76 × 10^−3^; OR = 0.97) (**Fig. 6A**). More specifically, the rs1905505-SNP is associated with decreased levels of whole blood glucose and HbA1C. While rs1905505 is not a known eQTL for SLC2A2, it does affect expression levels of genes located upstream that control islet responsiveness during metabolic stress: *RPL22L1*(ribosomal protein L22-like 1) and *EIF5A2* (eukaryotic translation initiation factor 5A2) (**Fig. 6B and 6C**)(Lonsdale et al. 2013; Raval et al. 2024; Maier et al. 2010). Visualization of the genomic landscape surrounding this islet-specific HMR highlights an enhancer-like signature with an overlapping islet DNA accessibility peak (**Fig. 6C and S7E**)(Varshney et al. 2017). As with the *GCK*-affiliated HMR sequences, we performed reporter assays for two different copies of the 219-bp DNA sequencing surrounding the rs1905505-SNP: one containing the T2D risk allele (A) and the other, the non-risk allele (G) (**Fig. 6D, Fig. S7F, and Table S15**). The risk allele rs1905505-A showed significantly decreased luciferase activity compared to the non-risk allele rs1905505-G in both INS1E and MIN6 cells (INS1E: p-value = 3.94 × 10^−4^; MIN6: p-value = 1.76 × 10^−8^). Enhancer activity represents an 8.8-fold (INS1E) to 13.6-fold (MIN6) decrease in transcriptional activity relative to the non-risk allele. Near rs1905505-SNP are other eQTLs, including rs1905504, rs7635100, and rs7635470, that affect *SLC2A2* gene expression in testis(Lonsdale et al. 2013). Together, they are all in LD in the European subpopulation of the 1000 Genome Phase 3 dataset (0.39 ≤ r^2^ ≤ 0.9). Accordingly, we examined transcriptional activity for a larger stretch of DNA (803-bp) inclusive of these SNP eQTLs (**Fig. 6E and Table S15**). We observed a decrease in transcriptional activity by 2.2-fold with the risk haplotype (A-C-T-C) in MIN6 cells (p-value = 5.25 × 10^−6^). However, results were not significant in INS1E cells (INS1E: p-value = 0.737). These findings demonstrate the transcriptional competence of this islet-specific HMR located upstream of *SLC2A2* as well as allele-specific enhancer activity of the rs1905505-SNP. In addition, cloning a DNA sequence with SNP eQTLs nearby rs1905505 did not confer an observable difference in the level of enhancer activity between non-risk and risk haplotypes (except for INS1E cells) compared to the shorter DNA sequences with just the non-risk and risk alleles for rs1905505 in either cell line. More importantly, our collective results using luciferase reporter assays for the *PAM*-, *GCK*- and *SLC2A2*-affiliated islet-specific HMRs demonstrate the utility of our research strategy for identifying islet-specific enhancers through integration of islet DNA methylation and genetic data.

**Figure 6.**
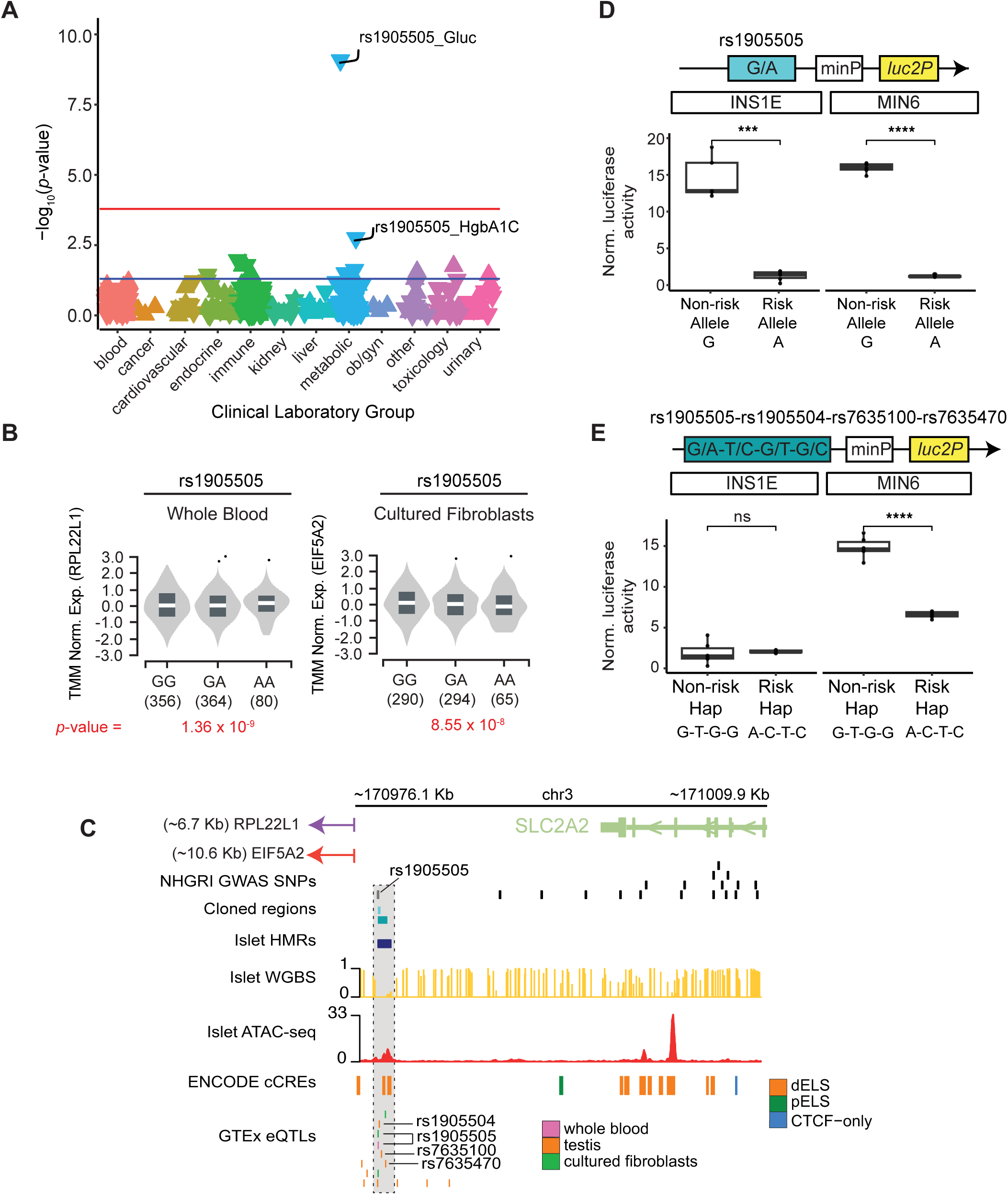
Islet-specific HMR with enhancer activity near *SLC2A2* harbors a T2D signal correlated with glucose levels. **(A)** Manhattan plot of laboratory-trait associations for T2D-associated SNP rs1905505. Each data point represents a laboratory-test association grouped and color-coded by clinical laboratory-test group on the x-axis and -log_10_(*p*-value) plotted on the y-axis. The orientation of the point indicates a positive (upward) or negative (downward) direction of effect on lab levels with each minor allele copy. The red and blue lines denote the Bonferroni-corrected (*p*-value < 1.62 × 10^−4^) and nominal (*p*-value = 0.05) significance level thresholds, respectively. **(B)** Multiple alignment of functional genomic data at the *SLC2A2*-associated islet-specific HMR containing SNPs highlighted in panel A. The islet WGBS track represents mean methylation levels at non-mutated CpG sites across islet donors, and the islet ATAC-seq data originates from the “Islet 1” sample described in Varshney et al. 2017(Varshney et al. 2017). (**C)** Violin plots showing normalized *RPL22L1* and *EIF5A2* expression associated with the rs1905505-eQTL in tissues from GTEx. Genotype counts and their associated sample sizes are displayed on the x-axis(Lonsdale et al. 2013). **(D)** Box plot representing functional validation of 219-bp segments flanking rs1905505. The risk allele (A) demonstrated significantly decreased transcriptional activity compared to the non-risk allele (G) in INS1E and MIN6 β-cells. Relative luciferase activity is given as a fold-change of Firefly:*Renilla* luciferase activity compared to the empty vector control. Significance was calculated using an unpaired two-sided *t*-test (n = 6 per haplotype)**. (E)** Box plot representing functional validation of 803-bp haplotype segments containing rs1905505, nearby *SLC2A2* eQTLs, and an ENCODE cCRE with a dELS. While no difference in transcriptional activity was observed in INS1E cells, the risk haplotype (A-C-T-C) showed significantly decreased enhancer activity than the non-risk haplotype (G-T-G-T) in MIN6 cells. Relative luciferase activity and *p*-values were determined using the same methods described in panel D (n=6 per haplotype); * = *p*-value < 0.05; ** = *p*-value < 0.01; *** = *p*-value < 0.001.

## DISCUSSION

Expanding T2D GWAS meta-analyses to include larger study cohorts has substantially increased the number of identified T2D risk loci(Vujkovic et al. 2020; Suzuki et al. 2024; Mahajan et al.). Recent integration of T2D GWAS and islet epigenomic data has revealed that T2D risk variants are significantly enriched in non-coding genomic regions, specifically islet enhancers linked to genes controlling β cell function and insulin secretion(Pasquali et al. 2014; Kycia et al. 2018; Varshney et al. 2017; Thurner et al. 2018; Greenwald et al. 2019; Cebola 2019). These studies argue that perturbed transcription at islet enhancers mediates a substantial portion of T2D risk, highlighting a critical need to elucidate key genetic regulatory mechanisms underlying islet dysfunction to better understand the pathophysiology of T2D.

Our previous work showed that DNA hypomethylation patterns at HMRs accurately identifies cell-type specific gene enhancers linked to cell-relevant biological pathways. We also demonstrated that HMRs serve as stable epigenetic markers of a cell’s developmental history, providing a reliable framework for identifying regulatory elements critical to cell-type identity and function(Gu et al. 2016; Roadmap Epigenomics Consortium et al. 2015; Schlesinger et al. 2013; Neri et al. 2017; Kleftogiannis et al. 2016; Benton et al. 2019). In the present study, we combined DNA methylation data from islets of four non-diabetic donors with genotype-phenotype data from BioVU, Vanderbilt’s DNA biobank, to identify gene enhancers associated with T2D risk. As a first step, we generated a reference set of ∼31,000 islet HMRs characterized by strong concordance in both methylation levels and genomic overlap across donors. Comparative methylation profiling of reference islet HMRs to WGBS data from other diverse cell types identified ∼4,800 islet-specific HMRs, the majority of which (≥ 50%) have enhancer-associated chromatin features, and are enriched for key islet TFs and associated genes essential for β cell development and function(Varshney et al. 2017). Of note, in addition to canonical islet TFs, we identified the neurogenesis-associated factor ATOH1 as significantly enriched in islet-specific HMRs. This finding points to potential similarities in their methylation profiles, consistent with existing evidence of shared transcriptomic features between the two cell types(Nica et al. 2013; Schmidt et al. 2024).

We further show that islet-specific HMRs are significantly enriched for GWAS SNPs linked to “Type 2 Diabetes” and “Hemoglobin A1C Levels” as reported in the NHGRI-EBI GWAS catalog(Sollis et al. 2023). Interestingly, traits not immediately attributable to islet function, including “Hematocrit”, “Prostate Cancer”, and “Serum Creatinine Levels” are also enriched in islet-specific HMRs, suggesting potential pleiotropic effects on gene expression in other organ systems. In line with these findings, S-LDSC confirmed islet-specific HMRs are enriched for the genetic contribution to heritability of “T2D” and “Glucose”, more so than the cell-type specific HMRs of other glucose-regulating tissues – adipose and liver. Overlapping islet-specific HMRs with T2D risk variants (MR-MEGA p-value = 5.0 × 10^−8^) from two major T2D GWAS meta-analyses underscored the functional relevance of the islet regulome to T2D risk(Zhong et al. 2010; De Silva et al. 2019). Across all cell types, islet-specific HMRs harbor significantly more T2D risk variants than a size-matched, randomized set of genomic regions. Although not directly involved in glucose regulation, B-cell and H1 ESC-specific HMRs also showed greater-than-expected overlap with T2D risk variants, albeit to a lesser extent than islet HMRs. While enhancer dysregulation in H1 ESCs has been linked to cancer, its connection to T2D risk remains unclear(Aran et al. 2016; Stergachis et al. 2013). Prior evidence of T2D variant enrichment in B-cells aligns with our S-LDSC and permutation test findings(Aylward et al. 2018).

With access to EHR-linked genetic data from the BioVU cohort of European ancestry (EU), we developed a region-based genetic association study, which we term “HMR-WAS”. The results of our HMR-WAS for T2D, which tested common SNPs (MAF > 1%) located in islet-specific HMRs, revealed a T2D-association signal (rs761773000: p-value = 3.54 × 10^−6^, OR = 1.28) found at a non-coding islet-specific HMR with demonstrated regulatory capacity surrounding the *PAM* gene locus. This gene encodes an enzyme that amidates proteins involved in granule exocytosis(Merkler et al. 2022). *PAM* is expressed at both the mRNA and protein levels in islet endocrine cells based on data from The Human Protein Atlas(Thul and Lindskog 2017). *PAM* mRNA levels are correlated with rs76177300 in thyroid and testis, but there is no evidence of PAM protein in either tissue(Lonsdale et al. 2013). These observations support our conclusion that we have identified an enhancer, which harbors this T2D-associated SNP, that may contribute to increased T2D risk by modulating *PAM* expression in islets specifically. Additionally, evidence linking *PAM* expression to this SNP in the cerebellum (where it is a called eQTL) highlights shared functional features between the endocrine and nervous system(Lkhagvasuren et al. 2021; Smith and Madson 1981).

It is important to note that variants in this *PAM* gene locus did not achieve Bonferroni- or genome-wide significance in a T2D GWAS of our BioVU EU cohort. This is likely attributable to power constraints and smaller effect sizes of the associated variants. This same locus, however, was strongly associated with T2D in our HMR-WAS, demonstrating the potential of our approach to uncover T2D-associated regulatory elements that are overlooked in genome-wide studies. Moreover, we expect this approach to be generalizable to other complex, polygenic traits.

Phenome-wide (PheWAS) and laboratory-wide (LabWAS) scans revealed that over 45% of T2D-associated, islet-specific HMRs contain at least one genetic association signal to a metabolic clinical phenotype or lab trait. Both analyses also suggest potential pleiotropic effects beyond metabolic traits like T2D, with secondary associations observed for dermatologic phenotypes and hematologic lab measures. Guided by our LabWAS findings, we prioritized two islet-specific HMRs with metabolic laboratory-trait associations that map to key regulators of glucose homeostasis: *GCK* and *SLC2A2*. These regions demonstrated enhancer activity and SNP haplotype-dependent effects on regulatory activity using luciferase reporter assays in both INS1E and MIN6 cells.

While INS1E and MIN6 are widely used rodent-derived islet β cell lines for studying β cell function, they may not fully capture the regulatory landscape of human β cells. However, due to shared orthologous TFs between human and rodent genomes, they remain useful for assessing candidate enhancer activity(Schmidt et al. 2010; Stergachis et al. 2014). Current human β cell line models, like the EndoC-βH1-3 cell lines, have their own limitations such as reduced insulin content (< 20%) and less mature β cell characteristics(Hastoy et al. 2018; Benazra et al. 2015). As human β cell lines improve, so will our ability to functionally characterize potential gene enhancers. Massively parallel reporter assays (MPRA) in future studies could enable broader validation of T2D-associated, islet-specific HMRs, extending the scope of regulatory element discovery.

Together, these findings highlight a framework for identifying T2D-associated, islet-specific regulatory regions. By integrating our reference islet methylation profile with clinical patient EHR, we prioritized islet-specific HMRs for functional validation *in vitro*. This approach confirmed the regulatory potential of three islet-specific HMRs associated with key genes involved in glucose metabolism: *PAM*, *GCK*, and *SLC2A2*. Altogether, this study presents a framework that integrates islet hypomethylation with patient genotype-phenotype data to identify islet-specific enhancers important to T2D risk, underscoring the importance of the islet regulome in T2D development and progression.

## METHODS

### Experimental model and study participant details

#### Islet procurement and processing

Pancreas procurement and processing, which includes islet isolation, was processed in Pittsburgh by R. Bottino as previously described(Brissova et al. 2018; Dai et al. 2017; Balamurugan et al. 2003). Islets were shipped to the Powers & Brissova Research Group at VUMC for further analysis following Integrated Islet Distribution Program (IIDP) shipping protocols. Genomic DNA (gDNA) from islet donor tissue (n = 4) was extracted using the Wizard Genomic DNA Purification Kit (Promega, #A1120). **Table S1** contains demographic information and clinical metrics of T2D on islet donors.

#### EHR-linked biobank BioVU at VUMC

The Vanderbilt Synthetic Derivative (SD) is a de-identified repository that includes patient EHR on more than 3 million patients at VUMC. In 2007, Vanderbilt launched a DNA biobank, called BioVU, that is linked to the SD. Today, it is one of the largest EHR-based biobanks in the world and has a collection of peripheral blood-derived DNA samples from than 300,000(Roden et al. 2008). Currently, >90,000 have been genotyped on the Illumina Expanded Multi-Ethnic Genotyping Array (MEGA^EX^). We obtained complete EHR and genome-wide genotype data from the Illumina MEGA^EX^ array on 28,807,316 genetic variants for 66,903 individuals of European ancestry (EU). The quality control process for samples and SNPs is described in detail in Dennis *et al*., 2021(Dennis et al. 2021).

As part of the HMR-WAS and GWAS for T2D in BioVU, we identified T2D cases and controls among BioVU individuals of European ancestry using the Northwestern University Type 2 diabetes mellitus algorithms. These algorithms apply stringent criteria to EMR data collected through routine clinical care, including diagnostic codes, medications, and lab test results, to more accurately classify individuals as true T2D cases or controls(Kho et al. 2012; Wei et al. 2012; Hripcsak et al. 2019; Pacheco and Thompson). Applying these algorithms refined the initial cohort of 56,064 BioVU EU subjects (9,749 cases and 46,315 controls) to a final analytic set of 17,404 consisting of 4,656 cases and 12,748 controls.

#### Cell lines

The MIN6 mouse insulinoma β cell line (a gift from and Kathy DelGiorno, VU), was grown in Dulbecco’s modified Eagle’s medium (Gibco, #11875135) supplemented with 110 mg/L sodium pyruvate, 4.5 g/L D-Glucose, 10% (v/v) heat-inactivated fetal bovine serum (FBS), 100 U/mL penicillin, 100 mg/mL streptomycin, and 0.1 mM β-mercaptoethanol. The INS-1E rat insulinoma β cell line (a gift from Maureen Gannon, VUMC) was cultured in RPMI 1640 culture medium (Gibco, #11875135) supplemented with 5% (v/v) heat-inactivated FBS, 1 mM sodium pyruvate, 50 µM 2-mercaptoethanol, 2 mM glutamine, 10 mM HEPES, 100 U/mL penicillin, and 100 µg/mL streptomycin. Both cell lines were passaged at 70-90% confluency with media changes every 2 days and maintained at 5% CO_2_, 37°C, and 80% humidity.

### Method details

#### Transposome preparation

The protocol for Tn5 transposase production and assembly was optimized and detailed previously(Barnett et al. 2020; Wang et al. 2013). To prepare the tagmentation-based WGBS (T-WGBS) transposome adapters, 10 μL each of oligonucleotides Tn5mC-Apt1 (100μM) and Tn5mC1.1-A1block (100μM) were combined with 80μL nuclease free water in a PCR tube. The adapters were annealed using a PCR thermocycler with the following program: 95°C for 3 minutes, 65°C for 3 minutes, ramp down to 24°C at a rate of - 1°C/second, hold at 24°C. Annealed oligos were then combined with 100μL glycerol to create a 5μM, 50% glycerol adapter mixture. Transposome assemblies were prepared by mixing equal parts of purified Tn5 transposase enzyme and annealed oligonucleotides, then incubating the mixture at 25°C for 60 min at room temperature. Oligonucleotides used for transposome assembly are provided in **Table S15**.

#### T-WGBS

Prior to T-WGBS library prep, the integrity of genomic DNA extracted from islet donor tissue was assessed using a Genomic DNA ScreenTape (Agilent, 5067-536) on an Agilent 2200 TapeStation. The four islet gDNA samples had an average DNA Integrity Number (DIN) of 9.2, reflecting high quality, intact gDNA suitable for library prep. T-WGBS libraries were prepared according to previously reported methods(Barnett et al. 2020; Wang et al. 2013; Adey and Shendure 2012). Genomic DNA was first diluted in a 50μL tagmentation reaction containing 80 ng gDNA, 2.5μL transposome, 10μL 5X Tris-DMF, and nuclease-free water to a final volume of 50μL. This reaction was incubated at 55°C for 8 minutes in a PCR thermocycler and then halted by adding 250μL Zymo DNA binding buffer from the DNA Clean & Concentrator-5 kit (Zymo, D4004). Column purification was carried out according to the manufacturer’s protocol instructions, and DNA was eluted in 15μL nuclease-free water at the final step.

A 1-2μL aliquot from each sample was reserved to evaluate fragmentation and size distribution post-tagmentation using a D5000 screentape (Agilent, 5067-5888) on an Agilent 2200 TapeStation. The remaining tagmented DNA (11μL DNA eluate) was used as input for the oligo replacement and gap repair reaction in a PCR tube consisting of 2μL 10μM Tn5mC-Repl01 oligonucleotide, 2μL 10x ampligase buffer (Lucigen, A3202K), and 2μL dNTP mix (2.5mM each). Gap repair was performed using the following PCR thermocycler program: 50°C for 1 minute, 45°C for 10 minutes, ramp down to 37°C at a rate of −0.1°C/second, hold at 37°C. Upon reaching 37°C, 1μL T4 DNA polymerase (NEB, M0203) and 2.5μL ampligase (Lucigen, A3202K) were added directly to the reaction tube without removal from the thermocycler. The reaction was gently mixed with repeated pipetting while minimizing bubble formation, then incubated at 37°C for 30 minutes, followed by at 4°C hold. Optionally, 2μL of the gap repair reaction was reserved for a test PCR to evaluate library distribution prior to bisulfite conversion. The remainder of the reaction was stopped by adding 102.5μL Zymo DNA binding buffer (5:1 buffer:DNA volume), then purified using the DNA Clean & Concentrator-5 kit (Zymo, D4004) and eluted in 20.5μL nuclease-free water.

For bisulfite conversion, 20.5μL gap-repaired DNA from the previous step was mixed with 130μL CT conversion reagent from the EZ DNA Methylation-Gold kit (Zymo, D5005). The mixture was split evenly across three PCR tubes (50 μL per tube) and incubated on a PCR thermocycler as follows: 98°C for 10 minutes, 64°C for 2.5 hours, hold at 4°C for up to 20 hours. After conversion, all three PCR tubes were pooled into a single tube, and in-column desulphonation and clean-up were performed according to the manufacturer’s protocol instructions. DNA was eluted in 25μl nuclease-water at final step.

Amplification and indexing of bisulfite-converted DNA was carried out in 50μL PCR reactions containing 25μL 2x KAPA HiFi HotStart Uracil+ ReadyMix (Roche, 07959052001), 20μL eluted bisulfite-converted DNA, and 1.5μL each of 10μM Nextera i5 and i7 index primers. PCR cycling parameters were: 98°C for 45 seconds, 10 cycles at 98°C for 15 seconds, 62°C for 30 seconds, 72°C for 30 seconds; followed by a final extension at 72°C for 2 minutes and a hold at 12°C. PCR-amplified DNA was purified using the DNA Clean and Concentrator-5 kit (Zymo, D4004) and eluted in 22μL nuclease free-water. Library quality and size distributions were evaluated using a D5000 screentape (Agilent, 5067-5888) on a Agilent 2200 TapeStation. Final T-WGBS DNA libraries were sequenced at the Vanderbilt VANTAGE core facility on the Illumina NovSeq 6000 using 2×150bp paired-end reads on, yielding approximately 170-270 million reads per library.

#### Dual luciferase reporter assay

##### Cloning test sequences into pGL4.27

We selected 15 DNA sequences from the *PAM*, *GCK*, and *SLC2A2* loci – five per locus – to evaluate for luciferase reporter activity (**Table S15**). All sequences, apart from the *SLC2A2* TSS-spanning sequence, overlap an islet-specific HMR. The islet-specific HMRs at these loci were selected based on their enhancer-like chromatin state and co-localization with genetic variants associated with T2D and/or clinical biomarkers of T2D(Varshney et al. 2017; Suzuki et al. 2024; Mahajan et al.). Their roles in insulin secretion and glucose regulation further reinforced our decision to investigate the regulatory potential of their associated islet-specific HMRs(Laukkanen et al. 2005; Fu et al. 2013). Sequences spanning the TSS of *GCK* (-249, +1), *PAM* (-249, +1) and *SLC2A2* (-113, +200) promoters were included as internal technical controls (**Fig. S7B, S7D, and S7F**).

All DNA fragments were designed with 40-bp overlapping ends required for Gibson cloning and synthesized by Integrated DNA Technologies (IDT). Each fragment’s 5’ and 3’ ends were appended with 40-bp sequences homologous to the left and right arms of the pGL4.27 vector, respectively:

- Left arm (5’→3’): TTTCTCTGGCCTAACTGGCCGGTACCTGAGCTCGCTAGCC
- Right arm (5’→3’): ATCAAGATCTGGCCTCGGCGGCCAAGCTTAGACACTAGAG

Each sequence was inserted into the multiple cloning site (MCS) of the pGL4.27 [luc2P/minP/Hygro] plasmid vector (Promega, E8451) with Gibson assembly. For cloning, the pGL4.27 vector backbone was linearized with a EcoRV-HF (NEB, R3195) and XhoI (NEB, R0416) double digest, followed by assembly using the NEBuider HiFi DNA Assembly Master Mix (NEB, E2621) at a 1:2 vector-to-insert ratio. Gibson products were transformed into NEB 5-alpha Competent *E. coli* (High Efficiency, NEB, C2987) cells following manufacturer guidelines. Plasmid DNA was purified using the ZR Plasmid Miniprep-Classic kit (Zymo, D4054), and clones were sequence-validated through Genewiz’s Plasmid-EZ sequencing service.

##### Dual-glo luciferase assay

Luciferase assays were conducted in two independent experiments (biological replicates), with six technical replicates per treatment condition for both INS-1E and MIN6 cell lines. Approximately 5.0 × 10^4^ INS-1E and 8.0 × 10^4^ cells MIN6 were seeded into 96-well plates. At 80% confluency, cells were co-transfected with luciferase reporter constructs (Firefly:*Renilla* plasmid ratio of 10:1) using Lipofectamine 2000 (Invitrogen, 11668) for INS-1E and Lipofectamine 3000 (Invitrogen, L30000) for MIN6 cells. For INS-1E cells, 0.15μg of pGL4.27 DNA (firefly luciferase plasmid with test insert) and 0.015μg pRL-TK DNA (*renilla* luciferase plasmid, Promega, E2241) were used; for MIN6 cells, 0.5 μg and 0.05μg of these plasmids, respectively. DNA-lipid complexes were prepared at 1:4 DNA:Lipofectamine ratio for INS-1E cells and 1:2 for MIN6. On each plate, we included non-transfected samples to control for background luminescence and GFP-transfected samples (pcDNA3.1-eGFP) to evaluate transfection efficiency. Forty-eight hours post-transfection, luciferase activity was measured using the Dual-Glo Luciferase Assay (Promega, E2920) per manufacturer guidelines. Firefly and *Renilla* luminescence were measured using a Promega Glo-Max Discovery luminometer with a 1-second integration time.

##### Luciferase reporter assay analysis

For each plate, the average firefly and *renilla* relative light unit (RLU) values from non-transfected samples served as background measurements for firefly and *Renilla* luciferase activity, respectively. Luminescence readings from samples transfected with firefly and *Renilla* constructs were background-corrected by subtracting these values. We then calculated firefly/*renilla* activity ratios by dividing the background-corrected firefly RLU value by the background-corrected *renilla* RLU value. These normalized luciferase activity values were used to determine fold-changes relative to the mean firefly/*renilla* activity ratio for samples that received the empty pGL4.27 vector control. Statistical significance between treatment groups was evaluated using an unpaired two-sided *t*-test with the *stat_compare_means* function from the *ggpubr* R package.

### Quantification and statistical analysis

#### T-WGBS library processing

All islet WGBS libraries from non-T2D donors were trimmed of adapters and evaluated for quality control using the Trim Galore! (version 0.6.6) Perl script wrapper for Cutadapt and FastQC(Martin 2011). WGBS reads were mapped with WALT (version 1.0) to the hg38 genome assembly, and preseq was used to compare library complexity across libraries(Chen et al. 2016; Daley and Smith 2013). DNA methylation analysis of mapped reads was performed using the *MethPipe* (version 5.0.1) software package, now *DNMTools*, which contains all existing *MethPipe* programs along with fixes and improvements. Methylation-specific statistics, such as library bisulfite conversion rate and single-site methylation levels at symmetric CpG sites, were generated after removing read duplicates(Song et al. 2013). Plots illustrating analysis statistics of donor islet HMR datasets, including distributions of HMR sizes, CpG count, and methylation levels, were generated using the *ggplot2* R package (**Fig. S1A-C**)(Hadley Wickham 2016). All code and specific parameters to process WBGS libraries can be found at: https://github.com/HodgesGenomicsLab/Islet_WGBS_T2D.

#### Reference islet HMR dataset

Using the *MethPipe* suite of tools, we generated files listing methylation levels at symmetric CpG sites (.meth) in each donor’s islet WGBS library using the *symmetric-cpgs* program with option “-m” to retain mutated sites. These methylation files were then used as input to the *hmr* program to identify HMRs in each donor. From the resulting HMR BED files, we excluded regions 51 bp or shorter to ensure a high-confidence HMR set for each donor(Song et al. 2013). We then assessed HMR overlap across individuals using the *intersect* function from the *Bedtools* package with option ‘-c’, which adds a column to each entry in the input ‘-a’ HMR BED file indicating the number of overlapping regions in the comparison ‘-b’ HMR BED files. We prioritized HMRs with a reported overlap count of three, which represent regions that are found in the ‘-a’ HMR BED file and all comparison ‘-b’ HMR BED files. HMRs that met this criterion from each donor were concatenated to generate a consensus HMR list, with overlapping regions consolidated using *bedtools merge* with default settings(Quinlan and Hall 2010). The resulting set of 35,721 HMRs constituted our reference islet HMR dataset. We used the *deeptools* suite of tools to analyze and visualize pairwise correlations between donors based on their CpG methylation profiles within reference islet HMRs. To assess overall similarity with cell types functionally unrelated to islets, we also compared each donor’s islet methylation profile to publicly available WGBS data from B cell and H1 ESC at the same set of regions (**Table S6, Fig. S1G**)(Ramírez et al. 2014; Song et al. 2013). Distributions of size, CpG count, and methylation levels for reference islet HMRs were visualized using the *ggplot2* R package (**Fig. S1D-F**). Reference methylation (.meth) and read coverage (.read) profiles for islets were generated by calculating averaging values at non-mutated symmetric CpG sites shared across all four donors using the *bedtools map* function with the ‘-o = mean’ option (n = 29,954,671)(Quinlan and Hall 2010). The resulting and read coverage profiles were visualized using UCSC’s function *bedGraphToBigWig* tool.

#### Comparative DNA methylation profiling

We relied on publicly available WGBS data to perform comparative DNA methylation profiling and obtained HMR datasets specifically through the *MethBase* trackhub, a central reference methylome database in the UCSC Genome Browser (**Table S6**). This UCSC trackhub consists of WGBS data processed through the *MethPipe* software package (now *DNMTools*)(Song et al. 2013). Among genome-wide methylation datasets with at least 10X coverage, we included cell types in crosstalk with islets to regulate blood sugar levels and collectively represent diverse organ systems. This resulted in the selection of: *H1 ESC* from Lister, et al.; *B cell* from Hodges, et al.; and *adipose* and *liver* from the NIH Roadmap Epigenomics Consortium(Lister et al. 2009; Hodges et al. 2011; Roadmap Epigenomics Consortium et al. 2015). As a primary cleaning step, we required greater than 51 bp in size for an HMR to be included in our comparative analysis. Next, we used the *intersect* function from the *Bedtools* package with option ‘-v’ to call ‘cell-type specific’ HMRs and option ‘-u’ to identify ‘shared’ HMRs, defined region with at least one base pair overlap as ‘shared’(Quinlan and Hall 2010).

#### Islet ChromHMM annotation

We obtained a 13-state ChromHMM annotation file for islets (hg19) from Varshney et al(Varshney et al. 2017). Following liftover of genomic coordinates from hg38 to hg19, we annotated islet-specific HMRs (n = 4,570), shared HMRs (n =30,821), and HMRs from the “islet-specific” *k*-means group (n = 10,265) based on their overlap with the islet chromatin states (**Fig. 2C and 2D**). Overlaps were evaluated using the *intersect* function from the *Bedtools* package with options ‘-wa’ and ‘-wb’, where the ‘-a’ file corresponded to one of the three islet HMR BED files and the ‘-b’ file was the islet ChromHMM annotation BED file(Quinlan and Hall 2010). Using R, we calculated the proportions of HMRs overlapping each ChromHMM state with denominators 6,232, 75,614, and 14,926 for islet-specific HMRs, shared HMRs, and “islet-specific” *k*-means HMRs, respectively. If an HMR overlapped multiple chromatin states, each overlapping state was included in the proportion calculations for its respective annotation. Bar plots visualizing annotation distributions were generated in R using the *ggplot2* package and ChromHMM state annotations are ordered by higher-level classification in the key to **Fig. 2D** as follows: transcribed (T), repressed (R), promoter (P), and enhancer (E).

#### Transcription factor motif enrichment analysis

Transcription factor (TF) motif enrichment analysis was performed on all cell-type specific HMR datasets (adipose, H1 ESC, B cell, islet, and liver) using the *HOMER* (version 4.10) software package (Table S7). BED files were scanned for enriched motifs using the *findMotifsGenome.pl* function with the option ‘-size given’ using randomly selected background regions generated by *HOMER* for comparison. Using output files listing known motif enrichment, we selected top representative TFs for each cell-type specific HMR dataset based on ranking by natural log-transformed binomial *p*-values. Scaled fold enrichment was calculated as the fold difference between two *HOMER* output values: *[%target/%background*](Heinz et al. 2010). Data was graphed in R using the *pheatmap* function from the *pheatmap* (version 1.0.12) package with options ‘scale = row’ and ‘cluster_rows *=* TRUE’(Kolde 2012). Enrichment values are scaled by row per TF motif to show relative cell-type specific enrichment.

#### Gene ontology enrichment analysis

Nearest neighbor gene annotation using the *annotatePeak* function in the ChIPseeker (version 1.30.3) package was used to identify genes uniquely associated for adipose-, H1 ESC-, B cell-, islet-, and liver-specific HMR datasets (**Table S7**)(Yu et al. 2015). Gene annotation parameters included defining the TSS region of a gene as 2 kb downstream and 1 kb upstream from the TSS(Yu et al. 2015). Gene ontology analysis was performed using the Reactome pathway database with the *compareCluster* function in the *clusterProfiler* (version 4.2.2) package to compare the functional roles of associated genes and their products across all cell-type specific HMR datasets. We selected top representative pathways for each cell-type specific HMR group based on ranking by FDR-adjusted *p*-values. Scaled fold enrichment was calculated as the quotient of two *clusterProfiler* output values: [*GeneRatio*/*BgRatio*](Wu et al. 2021). Graphing was performed in R using the *pheatmap* function from the *pheatmap* (version 1.0.12) package with options ‘scale = row’ and ‘cluster_rows = TRUE’(Kolde 2012). Enrichment values are scaled by row per pathway to show relative cell-type specific enrichment.

#### *k*-means clustering methylation heatmap

The methylation heatmap in **Fig. 2A** represents HMRs identified across all cell types (n_HMR_ = 89,882). To generate this consensus HMR set, we concatenated HMR BED files (size > 51 bp, **Table S7**) from each cell type and applied *bedtools merge* to vertically collapse overlapping regions(Quinlan and Hall 2010). To avoid potential methylation bias from non-autosomal, unlocalized or unplaced regions, we excluded HMRs located on chromosomes *chrUn*, *chrN_random*, *chrM*, *chrX* or *chrY*, reducing the region set from 94,393 to 90,205. We further filtered out HMRs lacking methylation data in any one cell type due to missing CpG methylation information, yielding a final list of 89,882 HMRs. CpG methylation profiles for adipose, B cell, liver, and H1 ESC were downloaded from the *MethBase* DNA methylation UCSC trackhub in the bigWig format and subsequently converted to bedGraph files (**Table S6**)(Hodges et al. 2011; Lister et al. 2009; Roadmap Epigenomics Consortium et al. 2015). For each cell type, we calculated the average methylation level for each region in the consensus HMR set using the *bedtools map* function with the ‘-o mean’ option, the consensus HMR BED file as the ‘-a’ file, and the corresponding cell type’s CpG methylation profile (.bedGraph) as the ‘-b’ file(Quinlan and Hall 2010). The resulting numerical matrix containing per-HMR methylation scores across all cell types was used as input for the *pheatmap* function from the pheatmap (version 1.0.12) package with options ‘scale = row’, ‘cluster_rows = TRUE’, and ‘kmeans_k = 6’(Kolde 2012). We used the elbow method to determine an optimal number of k-means clusters (**Fig. S2B**), selecting n = 6, and used the option “set.seed(2)” in R for reproducibility.

#### BioVU PheWAS

We performed PheWAS scans to better understand the phenotypic and potentially pleiotropic consequences of 163 T2D-associated SNPs (MR-MEGA *p*-value < 5.0 × 10^−8^) reported in the DIAMANTE and T2DGGI studies that localize to islet-specific HMRs and have a MAF > 1% in the BioVU EU cohort (**Fig. 3B**). Phenotypes in BioVU are mapped to phecodes, which represent custom groupings of diagnostic International Classification of Diseases (ICD-9/ICD-10) codes used to assign individuals a binary case or control status for a clinical disease or trait. Cases for each phecode were defined as having at least one relevant ICD-9 or ICD-10 code recorded on at least two distinct dates. Controls consisted of individuals who met the exclusion criteria for that phecode and had no diagnostic codes related to the phenotype of interest. A minimum of 20 cases per phecode was required for inclusion in our PheWAS analyses, resulting in the assessment of 1,749 phecodes in 66,278 BioVU individuals of European ancestry. These phecodes span 17 disease categories: infection diseases, neoplasms, endocrine/metabolic, hematopoietic, mental disorders, neurological, sense organs, circulatory system, respiratory, digestive, genitourinary, pregnancy complications, dermatologic, musculoskeletal, congenital anomalies, symptoms, and injuries/poisonings. Logistic regression was performed using the *phewas* function in the *PheWAS* R package (version 0.99.5-2) to identify phecodes that are significantly associated with genetic variation, assuming an additive genetic model. Analyses were adjusted for sex, median age and BMI across the medical record, and the top ten principal components of genetic ancestry to control for population stratification(Carroll et al. 2014; Denny et al. 2010, 2013). We applied a multiple testing correction for each SNP based on the number of valid association tests (i.e., those with non-missing *p*-values), using a Bonferroni threshold of 0.05 divided by the test count. This number varied slightly by SNP, ranging from 1,712 to 1,714. The code to run PheWAS in R is available on our GitHub page.

#### BioVU lab biomarker LabWAS

Unlike a PheWAS, which relies on phenotypes, a lab-wide association (LabWAS) study uses clinical laboratory (lab) measurements from the VUMC EHR to evaluate genetic associations with an independent variable. The lab quality control process is described in detail in Dennis *et al*., 2021, which identified 335 labs with non-zero heritability out of 471(Dennis et al. 2021). We prioritized heritable labs measured in at least 100 individuals, which resulted in testing 308 labs in 66,903 BioVU subjects of European ancestry. Labs are grouped into 12 subcategories: blood, metabolic, endocrine, kidney, immune, liver, urinary, OB/gyn, toxicology, cardiovascular and cancer. The same set of T2D-associated SNVs used in our PheWAS analyses (see BioVU PheWAS) were also tested for associations with lab traits. Linear regression was performed using the *lm()* function in the *stats* R package (version 4.1.0) to identify clinical labs that are significantly associated with genetic variation, assuming an additive genetic model. Analyses were adjusted for sex, median age and BMI across the medical record, and the top ten principal components of genetic ancestry. We calculated the Bonferroni-corrected statistical significance threshold for each SNP based on the total number of association tests performed (i.e., 0.05/308 = 1.62 × 10^−4^). The scripts to run LabWAS are available on our GitHub page.

#### GWAS and HMR-WAS for T2D

An HMR-WAS takes a targeted approach in contrast to a traditional GWAS that surveys the entire genome by evaluating variants within predefined genomic regions. As the name implies, association testing is limited to HMRs. Genomic coordinates of islet-specific HMRs (n_HMR_ = 4,676) were first lifted over from the hg38 assembly to hg19, yielding 4,570 HMRs. We excluded islet-specific HMRs on unplaced or unlocalized contigs, resulting in 4,511 regions mapped to autosomal or sex chromosomes. Using *PLINK* (version 2.0), we applied the “-maf 0.01” and “-extract” options to *PLINK*-generated files containing genotype (.bed), phenotype/pedigree (.fam) and variant (.bim) data from the BioVU T2D case-control cohort (n =17,404: 4,656 cases and 12,748 controls) to select only SNPs with MAF greater than 1% overlapping islet-specific HMRs. This resulted in 7,290 autosomal SNP tested for association with the T2D phecode (250.2). Logistic regression under an additive genetic model was performed in *PLINK*, adjusting for sex, median age and BMI across the medical record, and the top ten principal components of genetic ancestry(Purcell et al. 2007). A strong association signal was observed with rs76177300, prompting a follow-up HMR-WAS that included 313 additional SNPs in LD with rs76177300 (r^2^ ≥ 0.1, 1000kb). These SNPs were identified using genetic data from the CEU population (Utah Residents with Northern and Western European ancestry) in the 1000 Genomes Phase 3 dataset(Auton et al. 2015; Purcell et al. 2007). We conducted a T2D GWAS using the same BioVU T2D case-control cohort used in the T2D HMR-WAS. The T2D GWAS evaluated 5,523,377 SNPs (MAF > 1%) across the entire genome for association with T2D. Association results from the GWAS and HMR-WAS for T2D were visualized using Manhattan plots generated with the *ggplot2* R package. *P*-value distributions were evaluated using quantile-quantile (QQ) plots created with the *qqPlotFast* function in the *ramwas* R package (version 1.22.0). All *PLINK* commands and relevant code used in these analyses are available on our Github page.

#### S-LDSC

We quantified SNP-based genetic heritability across cell-type specific HMR datasets for adipose, H1 ESC, B cell, islets, and liver using stratified LD-score regression (S-LDSC). Reference base annotation files (Phase 3, version 2.2 annotations) aligned with the 1000 Genomes baseline v2.2 scores and HapMap 3 SNPs (https://alkesgroup.broadinstitute.org/LDSCORE/) were downloaded from the Price repository. Summary statistics for 84 clinical traits and laboratory measures (**Table S8**) were from the Price lab (https://alkesgroup.broadinstitute.org/LDSCORE/independent_sumstats/) and Neale lab heritability repository (https://nealelab.github.io/UKBB_ldsc/index.html) (Finucane et al. 2015). Traits were selected based on their relevance to general (cell-agnostic) disease pathophysiology or biological processes related to B cell, liver, or islet function. By including traits with diverse etiologies, we assessed the specificity of heritability enrichment for each cell-type specific HMR dataset. *S-LDSC* scripts were obtained from the Price lab GitHub (https://github.com/bulik/ldsc)(Bulik-Sullivan et al. 2015). The *S-LDSC* pipeline was run for each trait across all cell-type specific HMR datasets, and results were visualized using the *ggplot2* R package with functions *geom_point* and *case_when* for conditional coloring.

#### Trait enrichment analysis using the NHGRI-EBI GWAS Catalog

We downloaded version 1.0 of the NHGRI-EBI GWAS catalog (hg19) released on 03-09-2022 from the UCSC Genome Browser to perform trait enrichment tests across all cell-type specific HMR datasets for adipose, H1 ESC, B cell, islets, and liver (**Table S7 and Fig. 3A**)(Sollis et al. 2023). For each dataset, we excluded HMRs mapping to mitochondrial DNA (*chrM*) or an unknown chromosome (*chrUn*) to keep only HMRs located on chromosomes represented in the NGHRI-EBI GWAS catalog. We then used the *Bedtools intersect* function to identify overlapping SNPs and their associated traits using the HMR BED file as the ‘-a’ input and the NHGRI-EBI GWAS catalog (converted to BED format) as the ‘-b’ file(Quinlan and Hall 2010). The original NHGRI-EBI GWAS catalog was reformatted to keep only the following columns: ‘chrom’, ‘chromStart’, ‘chromEnd’, ‘name’, and ‘trait’. From the resulting SNP-trait overlaps, we performed a hypergeometric test to evaluate trait enrichment by comparing the frequency of each trait observed within a cell-type specific HMR dataset (i.e., sample) to its frequency in the GWAS catalog (i.e., population). Top representative traits for each cell-type specific HMR dataset were selected based on a hypergeometric *p*-value cut-og of 0.05. Scaled fold enrichment was calculated as the fold difference between trait frequency in the sample divided by the population(Sollis et al. 2023). Results were in R using the *pheatmap* function from the *pheatmap* (version 1.0.12) package with options ‘scale = row’ and ‘cluster_rows *=* TRUE’(Kolde 2012). The pipeline to run this analysis is available on our Github page.

#### Association analysis of genomics regions based on permutation testing

Multi-ancestry GWAS meta-analysis summary statistics were obtained from Mahajan et al. 2022 and Suzuki et al. 2024 (https://diagram-consortium.org/downloads.html). These studies reported 277 and 611 T2D risk loci, respectively. Notably, 275 loci identified in Mahajan et al. 2022 overlapped with loci reported in Suzuki et al. 2024. In total, 613 T2D risk loci were identified across both studies (Table S10)(Suzuki et al. 2024; Mahajan et al.). Due to differences in how T2D loci were defined and the specific SNVs tested for association, we utilized association summary statistics from both studies to maximize the overlap between islet-specific HMRs and T2D loci. Overlap analyses with reported T2D loci and summary statistics were conducted separately for each study, as detailed below. These results were then integrated to produce final overlap counts between islet-specific HMRs, T2D loci, and associated T2D signals, shown in **Fig. 3C**.

We began by identifying islet-specific HMRs that overlap T2D loci using *Bedtools intersect* with the option ‘-c’, where the ‘-a’ and ‘-b’ files were BED files listing islet-specific HMRs and T2D loci, respectively. This generated a BED file with an additional column indicating whether each HMR overlapped a T2D locus (1 for overlap, 1 for none). We further narrowed the subset of islet-specific HMRs overlapping T2D loci to those that also contained at least one genome-wide significant T2D-associated SNV (MR-MEGA *p*-value < 5.0 × 10^−8^). *Bedtools intersect* was run with the ‘-wa’ and ‘wb’ options, using HMRs marked with a ‘1’ from the previous intersect as the ‘-a’ file and genome-wide significant T2D-associated SNVs as the ‘-b’ file. The output was a BED file listing each instance of overlap between an HMR and a T2D-associated SNV. A similar approach was applied using *Bedtools intersect* to (1) identify T2D loci that overlap islet-specific HMRs and (2) further filter for T2D loci that overlap islet-specific HMRs containing at least one genome-wide significant T2D-associated SNV. In the first step, we used *Bedtools intersect* with the option ‘-c’, where the ‘-a’ and ‘-b’ files were T2D loci and islet-specific HMRs, respectively. In the second step, we used *Bedtools intersect* with the ‘-u’ option to identify T2D loci overlapping islet-specific HMRs that also contain at least one genome-wide significant T2D-associated SNV; the ‘-a’ file and ‘-b’ files corresponded to T2D loci and previously identified islet-specific HMRs containing genome-wide significant T2D signals, respectively(Quinlan and Hall 2010). For the overlap analysis using summary statistics from Suzuki et al. 2024, we used the common dbSNP153 catalog from the UCSC Genome Browser to assign RS numbers (rsID) to SNVs with genome-wide significant T2D association signals. This allowed us to identify which T2D-associated SNVs located at islet-specific HMRs were also present on the genotyping platform used for the BioVU European cohort (**Table S9**). This step was not necessary for the Mahajan et al. 2022 summary statistics because rsID numbers were provided.

We used the *regioneR* (version 1.40.1) R package to statistically evaluate and quantify the overlap between genome-wide significant T2D-associated SNVs (MR-MEGA *p*-value < 5.0 × 10^−8^) identified across both DIAMANTE and TDGGI studies (n_T2D SNV_ = 83,690) and cell-type specific HMR datasets for adipose, H1 ESC, B cell, islets, and liver (**Table S7 and Fig. 3D**)(Gel et al. 2016). Overlap testing was performed using the *overlapPermTest* function, where the ‘A’ file corresponded to a *GRanges* object of an HMR BED file and the “B” file, a *GRanges* object listing T2D-associated SNVs. The test was run using the following options with the hg19 *BSgenome* assembly loaded into the environment: ‘alternative = greater’ and ‘ntimes = 10000’. Although typically used for reproducibility, we did not apply *set.seed()* or the ‘mc.set.seed’ option in the *overlapPermTest* function as doing so introduced issues affecting the generation of randomized regions used for overlap comparison across all cell types. The permutation test results for liver shown in Fig. 3D were based on a representative subset of 6,329 liver-specific HMRs randomly sampled from the full set of 16,284 using *bedtools sample*(Quinlan and Hall 2010). This down-sampling was done to better align the liver region count with those of the other cell types (islet: 4,570, H1 ESC: 4,696, B cell: 7,999, and adipose: 7,748). The sample size of 6,239 corresponds to the mean of these four datasets.

#### Visualization and figure creation

Images and figures were generated using ggplot2 (version 3.5.1), plotgardener (1.0.17), deeptools (2.29.2), pheatmap (1.0.12), and eulerr (7.0.0)(Kramer et al. 2022; Kolde 2012; Micallef and Rodgers 2014; Hadley Wickham 2016). Schematic of study pipeline was created using Biorender.com.

## Data Availability

All data produced in the present study have been deposited in the Gene Expression Omnibus (GEO), but the GEO repository (GSE302385) is currently private. As such, all data produced in this study are available upon reasonable request to the authors.

## DATA ACCESS

All islet WGBS data have been deposited in the Gene Expression Omnibus: WGBS (GEO: GSE302385). The islet ATAC-seq data originates from sample “Islet 1” from the publication Varshney et al., 2017. GTEx eQTL data for genes *PAM*, *SLC2A2*, and *GCK* were obtained from the GTEx portal. Software and packages used are listed in **Table S16**. Detailed workflows and all code used to conduct the analyses in this paper are publicly available at: https://github.com/HodgesGenomicsLab/Islet_WGBS_T2D.

## ETHICS APPROVAL AND CONSENT TO PARTICIPATE

The Vanderbilt University Medical Center (VUMC) Institutional Review Board (IRB) oversees use of the Vanderbilt BioVU patient data repository and gave ethical approval for this project (IRB# 190418). Human islets used in this study were obtained from organ donors through the Integrated Islet Distribution Program (IIDP). De-identified donor information, medical histories and clinical characteristics were correlated with data generated in this study. The Vanderbilt University Institutional Review Board declared that studies on de-identified human pancreatic specimens are exempt from human subject studies approval.

## ACKNOWLEDGEMENTS

The authors thank the organ donors and their families, without whom this work would not be possible. The BioVU biobank was launched by VUMC in 2007 and consists of individuals who receive care at VUMC and are willing to participate in the BioVU research study. A patient’s DNA sample was linked with their electronic health records, including data on billing codes and clinical laboratory test results. Informed consent has been obtained from all patients involved in the study. Sequencing and data analysis were supported by resources from the Vanderbilt Advanced Computer Center for Research and Education (ACCRE) and Vanderbilt Technologies for Advanced Genomics (VANTAGE). We are grateful for support of the project by NIH awards (R01 GM147078 to E.H., R01 DK129469, U01DK135017 to M.B.), Department of Defense Idea award (W81XWH-20-1-0522 to E.H.), an American Cancer Society (ACS) Institutional Research Grant (#IRG-15-169-56 to E.H.), the Vanderbilt University Stanley Cohen Innovation Fund (to E.H), the VU School of Medicine Dean’s Faculty Fellow Award (to E.H) and funds from the Vanderbilt Ingram Cancer Center. Human pancreatic islets and/or other resources were provided by the NIDDK-funded IIDP (RRID:SCR_014387) at City of Hope, NIH Grant # U24DK098085.

## Author Contributions

VM and EH conceived of the project. VM generated the islet WGBS libraries and conducted the computational experiments and data analysis. VM cultured the cells and performed the luciferase reporter assay experiments. TS contributed to the S-LDSC analysis. ED developed the computational pipeline for trait enrichment analysis. AXM contributed to experimental design. MB and AP aided collaboration to collect genomic DNA samples from human islet donor tissue. SM extracted islet genomic DNA from human islet donor tissue. VM and EH wrote and edited the manuscript and figures. All authors reviewed and approved the final manuscript.

