## Supplementary Tables and Figures for "Islet-specific DNA hypomethylation identifies gene enhancer loci driving Type 2 Diabetes risk"

### SUPPLEMENTAL TABLE LEGENDS

**Table S1. Characteristics of non-T2D islet donors included in the WGBS analysis.** This table summarizes donor demographic information (age, sex, and genetic ancestry) and clinical metrics of T2D (BMI and HbA1C levels).

**Table S2. Sequencing and analysis statistics for islet WGBS libraries from each non-T2D donor.** This table contains statistics related to mapping and CpG sites obtained through the MethPipe software package for each donor's islet WGBS library.

**Table S3. Islet HMRs categorized by consensus category for each non-T2D donor after size selection.** This table summarizes findings from a comparison of donor islet HMR profiles. After size selection, each donor's islet HMRs was classified into four consensus categories based on their overlap with HMRs in the other individuals. These categories were subsequently used to sort HMRs into our curated reference and non-reference islet HMR datasets.

**Table S4. Mean coverage values for CpGs in islet HMRs by consensus category for each non-T2D donor.** This table summarizes the mean and standard deviation of CpG site coverage within islet HMRs stratified by consensus category. Refer to **Table S3** for a description of the categories.

**Table S5. Coverage statistics for non-mutated symmetric CpG sites found in our reference islet methylation profile.** This table reports statistics related to CpG sites in the reference islet methylation profile, including total CpG count, number of sites with at least 5X read coverage, and average read depth.

**Table S6. Coverage values for WGBS datasets of cell types used for comparative methylation profiling.** This table summarizes details on the publicly available WGBS datasets used in our comparative WGBS analysis (H1 ESC, B cell, liver, and adipose cells), including their sources and average CpG site coverage.

**Table S7. Number of shared and cell-type specific HMRs per cell type based on comparative methylation profiling.** This table summarizes findings from comparative methylation profiling of HMR datasets from islets, H1 ESC, B cell, liver, and adipose cells. After size selection, each cell type's HMRs was classified into two categories – cell-type specific and shared – based on their overlap with HMRs in the other cell types.

**Table S8. List of 84 summary statistic files used for S-LDSC analyses.** This table lists the names of traits and clinical values for which GWAS summary statistics were obtained from the Alkes Price lab website or Neale lab summary statistics browser.

**Table S9. Summary statistics for T2D-associated SNVs from the DIAMANTE and T2DGGI studies found in islet-specific HMRs.** This table lists summary statistics and locus details for a subset of SNVs (n = 255) evaluated for T2D association in the multi-ancestry GWAS meta-analyses conducted by the DIAMANTE and T2DDGI studies. These SNVs were specifically prioritized for their co-localization with islet-specific HMRs and their genome-wide significant T2D signals (MR-MEGA $p$-value < $5.0 \times 10^{-8}$) in one or both studies. An additional column indicates whether each T2D-associated SNV has corresponding genetic data with a MAF of at least 1% in the BioVU EU population.

**Table S10. Genomic overlap analysis between islet-specific HMRs and T2D loci from the DIAMANTE and T2DGGI studies.** This table lists locus details on 613 T2D loci identified across both studies along with any overlapping islet-specific HMRs.

**Table S11. Summary statistics of phenotypic associations for T2D-associated SNPs found in islet-specific HMRs using BioVU records.** This table lists summary statistics of phenotypic associations for T2D-associated SNPs located within islet-specific HMRs and have corresponding genetic data with MAF > 1% in the BioVU EU population (see **Table S9**). This resulted in 163 SNPs evaluated for association with 1,749 phecodes using BioVU records for 66,278 individuals of European descent.

**Table S12. Summary statistics of laboratory-trait associations for T2D-associated SNPs found in islet-specific HMRs using BioVU records.** This table lists summary statistics of laboratory-trait associations for T2D-associated SNPs located within islet-specific HMRs and have corresponding genetic data with MAF > 1% in the BioVU EU population (see **Table S9**). This resulted in 163 SNVs evaluated for association with 308 labs using BioVU records for 66,903 individuals of European descent.

**Table S13. Summary statistics from HMR-WAS for T2D using BioVU records.** This table presents summary statistics from an HMR-WAS for T2D using genotype-phenotype data from 17,404 individuals of European ancestry (EU) in BioVU (4,656 cases and 12,748 controls). The analysis included 7,290 SNPs on the Illumina $MEGA^{EX}$ array that are located within islet-specific HMRs and have MAF > 1% in the BioVU EU population ($N$ = 66,278). To more comprehensively characterize a strong association signal at rs76177300 found on chromosome 5 (see **Fig. 4B**), additional SNPs in LD with this variant (1000 kb window, $r^2 \geq 0.1$) were incorporated, bringing the total number of SNPs tested to 7,603.

**Table S14. Summary statistics from GWAS for T2D using BioVU records.** This table presents summary statistics from a GWAS for T2D using genotype-phenotype data from 17,404 individuals of European ancestry (EU) in BioVU (4,656 cases and 12,748 controls). The analysis included 5,523,377 SNPs on the Illumina $MEGA^{EX}$ array that have MAF > 1% in the BioVU EU population ($N$ = 66,278).

**Table S15. DNA sequences cloned into luciferase reporter plasmid.** This table provides details on the 15 DNA sequences spanning the *PAM*, *GCK*, and *SLC2A2* loci that were cloned into the pGL4.27 firefly luciferase vector.

**Table S16. Key resources table.** This table lists details on key resources used for this study, including commercial assays, experimental models, and software.

### SUPPLEMENTARY TABLES

**Supplemental Table 1. Characteristics of non-T2D islet donors included in the WGBS analysis.**

| | **Donors without T2D (*n* = 4)**[a] |
|---|---|
| **Sex (M/F)** | 3/1 |
| **Age (years)** | 46.5 ± 7.5 (31-63) |
| **BMI (kg/m²)**[b] | 26.3 ± 0.72 (24.7-28.0) |
| **HbA1C (%)**[b] | 5.35 ± 0.16 (4.9 - 5.6) |
| **HbA1C (mmol/mol)**[b] | 37 ± 1.70 (32 - 39) |
| **Race, n (%)** | |
| **White** | 2 (50%) |
| **African American** | 2 (50%) |

[a] Data are presented as mean ± SEM (range).

[b]These measurements were taken at the time of each donor's final hospitalization.

BMI: body mass index.

HbA1C: hemoglobin A1c.

**Supplemental Table 2. Sequencing and analysis statistics for islet WGBS libraries from each non-T2D donor.**

| **Donor ID** | **Total read pairs** | **Uniquely mapped reads** | **Total CpGs (# non-mutated; # mutated)** | **CpGs coverage ≥ 5X** | **CpGs mean depth covered** |
|---|---|---|---|---|---|
| Sample 1 | 173,742,827 | 113,019,491 | 30,961,395 (30,601,877; 359,518) | 17,658,412 | 5.29 |
| Sample 2 | 271,840,365 | 189,905,025 | 30,963,210 (30,563,854; 399,356) | 23,006,154 | 7.75 |
| Sample 3 | 262,471,678 | 182,959,629 | 30,956,395 (30,585,239; 371,156) | 22,690,825 | 7.44 |
| Sample 4 | 249,023,149 | 173,544,964 | 30,960,330 (30,552,252; 408,078) | 21,720,435 | 6.81 |

**Supplemental Table 3. Islet HMRs categorized by consensus category for each non-T2D donor after size selection.**

| **Donor ID** | **# of HMRs** | **# of HMRs > 51 bp** | **Consensus Category** | | | |
|---|---|---|---|---|---|---|
| | | | **3**[a] | **2**[b] | **1**[c] | **Unique**[d] |
| Sample 1 | 45,599 | 45,556 | 36,370 | 3,874 | 2,563 | 2,749 |
| Sample 2 | 50,799 | 50,676 | 36,763 | 5,874 | 3,988 | 4,051 |
| Sample 3 | 50,936 | 50,823 | 36,610 | 6,075 | 4,235 | 3,903 |
| Sample 4 | 51,199 | 51,082 | 37,196 | 5,675 | 3,860 | 4,351 |
| **# of reference islet HMRs:** | | | 35,721[e] | | | |

| # of non-reference islet HMRs: | 30,061[f] |
|---|---|

[a]HMRs(> 51 bp) assigned to consensus category '3' physically overlap HMRs (by at least 1 bp) found in all donors being compared to for that individual.

[b]HMRs (> 51 bp) assigned to consensus category '2' physically overlap HMRs (by at least 1 bp) found in any two of the other three donors being compared to for that individual.

[c]HMRs (> 51 bp) assigned to consensus category '1' physically overlap HMRs (by at least 1 bp) found in any one of the other three donors being compared to for that individual.

[d]HMRs (> 51 bp) assigned to consensus category 'Unique' are donor-specific and do not overlap HMRs found in the other donors being compared to for that individual.

[e]HMRs assigned to consensus category '3' across all donors were concatenated and vertically collapsed to merge any overlapping regions to curate a final list of reference islet HMRs.

[f]HMRs assigned to consensus categories '2', '1', or 'Unique' across all donors were concatenated and vertically collapsed to merge any overlapping regions to curate a final list of non-reference islet HMRs.

**Supplemental Table 4. Mean coverage values for CpGs in islet HMRs by consensus category for each non-T2D donor.**

| Donor ID | Consensus Category | | | | SD |
|---|---|---|---|---|---|
| | 3 | 2 | 1 | Unique | |
| Sample 1 | 5.68 | 5.84 | 5.40 | 4.72 | 0.49 |
| Sample 2 | 8.69 | 9.11 | 8.01 | 6.71 | 1.05 |
| Sample 3 | 8.38 | 8.67 | 7.96 | 6.74 | 0.85 |
| Sample 4 | 7.67 | 7.89 | 7.40 | 6.73 | 0.50 |

SD = standard deviation.

**Supplemental Table 5. Coverage statistics for non-mutated symmetric CpG sites found in our reference islet methylation profile.**

| Total CpGs | CpGs coverage ≥ 5X | CpGs mean depth covered |
|---|---|---|
| 29,954,671 | 22,508,813 | 6.82 |

**Supplemental Table 6. Coverage values for WGBS datasets of cell types used for comparative methylation profiling.**

| Cell type | Source | Genome assembly | Coverage | Download link |
|---|---|---|---|---|
| H1 ESC | Widespread Differences in Human DNA | GRCh38/hg38 | 25.933 | https://hgdownload.soe.ucsc.edu/hubs/methbase/v1/h |

| | | | | |
|---|---|---|---|---|
| | Methylomes, Lister 2009 | | | g38/Human_H1ESC.hmr.bb |
| B cell | Changes in Human Hematopoietic Stem Cells, Hodges 2011 | GRCh38/hg38 | 11.855 | https://hgdownload.soe.ucsc.edu/hubs/methbase/v1/hg38/Human_BCell.hmr.bb |
| Liver | Roadmap 2015 | GRCh38/hg38 | 49.619 | https://hgdownload.soe.ucsc.edu/hubs/methbase/v1/hg38/Human_Liver.hmr.bb |
| Adipose cells | Roadmap 2015 | GRCh38/hg38 | 129.245 | https://hgdownload.soe.ucsc.edu/hubs/methbase/v1/hg38/Human_Liver.hmr.bb |

**Supplemental Table 7. Number of shared and cell-type specific HMRs per cell type based on comparative methylation profiling.**

| Cell type | Total HMRs | HMRs > 51 bp | Cell type-specific[a] | Shared[b] |
|---|---|---|---|---|
| Adipose | 58,898 | 58,786 | 7,755 | 51,031 |
| Liver | 58,544 | 58,445 | 16,294 | 42,151 |
| H1 ESC | 36,322 | 35,914 | 4,697 | 31,217 |
| B-cell | 52,836 | 52,686 | 8,004 | 44,682 |
| Islet | 35,721 | 35,721 | 4,858 | 30,863 |

[a]Among HMRs > 51 bp, cell-type specific HMRs do not physically overlap HMRs found in the other cell types being compared to for that cell type.

[b]Among HMRs > 51 bp, shared HMRs physically overlap (by at least 1 bp) HMRs found in at least one of the four other cell types being compared to for that cell type.

**Supplemental Table 8. List of 84 summary statistic files used for S-LDSC analyses.**

| Trait[a] |
|---|
| Albumin |
| ALP |
| ALT |
| Angina_byDoctor |
| Apolipoprotein_B |
| AST |
| blood_EOSINOPHIL_COUNT |

| |
|---|
| blood_PLATELET_COUNT |
| blood_RBC_DISTRIB_WIDTH |
| blood_RED_COUNT |
| blood_WHITE_COUNT |
| bmd_HEEL_TSCOREz |
| body_BALDING1 |
| body_BMIz |
| body_HEIGHTz |
| body_WHRadjBMIz |

[a]Files were sourced from the Alkes Price lab website or Neale lab summary statistics browser.

**This is a sample table of Supplemental Table 8. The complete table is included as a separate Excel file.**

**Supplemental Table 9. Summary statistics for T2D-associated SNVs from the DIAMANTE and T2DGGI studies found in islet-specific HMRs.**

| Islet-specific HMR coordinates (bp, b37) | SNV[a] | SNV position (bp, b37) | Genotype data available in BioVU EU cohort (MAF > 1%) | Alleles | |
|---|---|---|---|---|---|
| | | | | Risk | Other |
| chr11:2857659-2858697 | rs2237897 | chr11:2858545-2858546 | no | T | C |
| chr11:2857659-2858697 | rs2237896 | chr11:2858439-2858440 | no | A | G |
| chr11:2857659-2858697 | rs74046911 | chr11:2858635-2858636 | no | T | C |
| chr11:2857659-2858697 | rs2299620 | chr11:2858294-2858295 | no | T | C |
| chr17:36101191-36104285 | rs11651052 | chr17:36102380-36102381 | no | A | G |
| chr17:36101191-36104285 | rs11263763 | chr17:36103564-36103565 | no | A | G |
| chr17:36101191-36104285 | rs8064454 | chr17:36101585-36101586 | no | A | C |

[a]T2D-associated SNV maps to an islet-specific HMR.

SNV: Single nucleotide variant.

**This is a sample table of Supplemental Table 9 that doesn’t show all columns. The complete table is included as a separate Excel file.**

**Supplemental Table 10. Genomic overlap analysis between islet-specific HMRs and T2D loci from the DIAMANTE and T2DGGI studies.**

| Suzuki et al., 2024 (T2DGGI) | | Mahajan et al., 2022 (DIAMANTE) | | Number of overlapping DIAMANTE loci[c] | Number of overlapping islet-specific HMRs |
|---|---|---|---|---|---|
| Locus[a] | Locus interval (bp, b37) | Locus[b] | Locus interval (bp, b37) | | |
| GINS2 | chr16:85204584-86959737 | NA | NA | NA | 33 |
| FAM101A, NCOR2 | chr12:124008976-125326462 | MPHOSPH9-ZNF664 | chr12:123118544-125045435 | 1 | 26 |
| VPS53 | chr17:19554-2809929 | NA | NA | NA | 22 |
| FEN1 | chr11:60745614-62065908 | NA | NA | NA | 21 |
| TCF3 | chr19:318284-2365673 | NA | NA | NA | 21 |

[a]A cell with 'NA' represents a T2D risk locus reported in Mahajan et al. 2022 but not Suzuki et al. 2024.

[b]A cell with 'NA' represents a T2D risk locus reported in Suzuki et al. 2024 but not Mahajan et al. 2022.

[c]This column shows the number of T2D risk loci reported in Mahajan et al. 2022 that overlap the boundaries of a corresponding T2D risk locus identified in Suzuki et al. 2024. The associated overlapping loci are listed in the third column. A cell with 'NA' indicates no overlap, meaning the associated T2D risk locus was uniquely identified in Suzuki et al. 2024.

**This is a sample table of Supplemental Table 10 that doesn't show all columns. The complete table is included as a separate Excel file.**

**Supplemental Table 11. Summary statistics of phenotypic associations for T2D-associated SNPs found in islet-specific HMRs using BioVU records.**

| T2D-associated SNP[a] | Phenotype | Phenotype description | Phenotype group | HWE P | UNADJ P | OR |
|---|---|---|---|---|---|---|
| rs2523504_T | 335 | Multiple sclerosis | neurological | 0.986 | 1.25E-10 | 1.331 |
| rs2239525_G | 335 | Multiple sclerosis | neurological | 0.987 | 1.54E-10 | 1.330 |
| rs2239526_G | 335 | Multiple sclerosis | neurological | 0.987 | 1.54E-10 | 1.330 |
| rs2239525_G | 250.1 | Type 1 diabetes | endocrine/metabolic | 0.984 | 1.54E-10 | 0.788 |
| rs2239526_G | 250.1 | Type 1 diabetes | endocrine/metabolic | 0.984 | 1.54E-10 | 0.788 |
| rs2523507_G | 250.1 | Type 1 diabetes | endocrine/metabolic | 0.983 | 1.61E-10 | 0.788 |
| rs2523507_G | 335 | Multiple sclerosis | neurological | 0.988 | 2.27E-10 | 1.326 |

| | | | | | | |
|---|---|---|---|---|---|---|
| rs2523504_T | 250.1 | Type 1 diabetes | endocrine/metabolic | 0.982 | 2.71E-10 | 0.791 |
| rs2239525_G | 250 | Diabetes mellitus | endocrine/metabolic | 0.986 | 1.42E-07 | 0.904 |

[a]T2D-associated SNP: SNP with a genome-wide significant T2D signal (as measured in one or both DIAMANTE and T2DGGI studies) that has MAF > 1% and corresponding genetic data in individuals of European ancestry in BioVU.

T2D-associated SNP: Variant identifier.

HWE P: The Hardy-Weinberg equilibrium p-value for the predictor.

UNADJ P: Unadjusted p-value.

OR: Odds ratio.

**<u>This is a sample table of Supplemental Table 11 that doesn’t show all columns. The complete table is included as a separate Excel file.</u>**

**Supplemental Table 12. Summary statistics of laboratory-trait associations for T2D-associated SNPs found in islet-specific HMRs using BioVU records.**

| T2D-associated SNP[a] | Lab | Full name of lab | Group | UNADJ P | OR |
|---|---|---|---|---|---|
| rs4400674_A | MCH | MCH [Entitic mass] by Automated count | blood | 1.95E-12 | 1.036 |
| rs7693_T | MBRat | Creatine kinase.MB/Creatine kinase.total in Serum or Plasma | cardiovascular | 8.06E-10 | 1.082 |
| rs7632381_C | MCV | MCV [Entitic volume] by Automated count | blood | 1.08E-08 | 0.972 |
| rs2403205_T | MBRat | Creatine kinase.MB/Creatine kinase.total in Serum or Plasma | cardiovascular | 1.44E-08 | 1.075 |
| rs724016_G | MCV | MCV [Entitic volume] by Automated count | blood | 1.45E-08 | 0.972 |
| rs76369685_A | Gluc | Glucose lab | metabolic | 3.63E-08 | 0.941 |
| rs79896666_C | Gluc | Glucose lab | metabolic | 3.64E-08 | 0.941 |
| rs78551082_T | Gluc | Glucose lab | metabolic | 3.64E-08 | 0.941 |
| rs74628648_T | Gluc | Glucose lab | metabolic | 3.64E-08 | 0.941 |

[a]T2D-associated SNP: SNP with a genome-wide significant T2D signal (as measured in one or both DIAMANTE and T2DGGI studies) that has MAF > 1% and corresponding genetic data in individuals of European ancestry in BioVU.

T2D-associated SNP: Variant identifier.

UNADJ P: Unadjusted p-value.

OR: Odds ratio.

**This is a sample table of Supplemental Table 12 that doesn't show all columns. The complete table is included as a separate Excel file.**

**Supplemental Table 13. Summary statistics from HMR-WAS for T2D using BioVU records.**

| Islet-specific HMR coordinates (bp, b37)[a] | SNP | SNP position (bp, b37) | in LD with rs76177300 | Alleles | | OR | SE | UNADJ P |
|---|---|---|---|---|---|---|---|---|
| | | | | Risk | Other | | | |
| NA | rs114964731 | chr5:102069652-102069653 | 1 | A | C | 1.40 | 0.07 | 1.57E-06 |
| NA | rs78761111 | chr5:102072478-102072479 | 1 | C | T | 1.38 | 0.07 | 1.91E-06 |
| NA | rs75457267 | chr5:102658769-102658770 | 1 | T | C | 1.38 | 0.07 | 3.19E-06 |
| NA | rs78162670 | chr5:101662385-101662386 | 1 | G | A | 1.38 | 0.07 | 3.24E-06 |
| NA | rs77487268 | chr5:101675581-101675582 | 1 | A | G | 1.38 | 0.07 | 3.25E-06 |
| NA | rs116782923 | chr5:102331464-102331465 | 1 | T | A | 1.36 | 0.07 | 3.49E-06 |
| chr5:102143245-102145396 | rs76177300 | chr5:102143310-102143311 | 1 | A | G | 1.40 | 0.07 | 3.54E-06 |
| NA | rs79602013 | chr5:101779345-101779346 | 1 | C | T | 1.40 | 0.07 | 3.58E-06 |
| NA | rs114555443 | chr5:102128957-102128958 | 1 | C | T | 1.38 | 0.07 | 4.50E-06 |

[a]SNPs with 'NA' for islet-specific HMR coordinates do not reside in an islet-specific HMR. Rather, they are part of the cluster of SNPs in LD with rs76177300 (1000 kb window, $r2 \geq 0.1$) based on CEU population data from 1000 Genomes Phase 3.

SNP: Variant identifier.

in LD with rs76177300: Binary indicator (1/0) categorizing SNPs within 1000 kb of rs76177300 and in LD at $r2 \geq 0.1$ based on CEU population data from Phase 3 of the 1000 Genomes Project.

OR: Odds ratio.

SE: Standard error of odds ratio estimate.

UNADJ P: Unadjusted p-value.

**This is a sample table of Supplemental Table 13 that doesn't show all columns. The complete table is included as a separate Excel file.**

**Supplemental Table 14. Summary statistics from GWAS for T2D using BioVU records.**

| SNP[a] | SNP position (bp, b37) | Allele | | OR | SE | UNADJ P | BONF | FDR |
|---|---|---|---|---|---|---|---|---|
| | | Risk | Other | | | | | |
| rs34872471 | chr10:114754070-114754071 | C | T | 1.325 | 0.032 | 1.02E-18 | TRUE | TRUE |
| rs35198068 | chr10:114754783-114754784 | C | T | 1.326 | 0.032 | 1.04E-18 | TRUE | TRUE |
| rs7903146 | chr10:114758348-114758349 | T | C | 1.325 | 0.032 | 1.36E-18 | TRUE | TRUE |
| rs4506565 | chr10:114756040-114756041 | T | A | 1.305 | 0.031 | 1.95E-17 | TRUE | TRUE |
| rs7901695 | chr10:114754087-114754088 | C | T | 1.301 | 0.031 | 4.58E-17 | TRUE | TRUE |
| rs4575195 | chr10:114765746-114765747 | A | C | 1.297 | 0.031 | 1.20E-16 | TRUE | TRUE |
| rs4132670 | chr10:114767770-114767771 | A | G | 1.296 | 0.031 | 2.11E-16 | TRUE | TRUE |
| rs36090025 | chr10:114774432-114774433 | C | A | 1.298 | 0.032 | 2.46E-16 | TRUE | TRUE |
| rs7074440 | chr10:114785423-114785424 | A | G | 1.298 | 0.032 | 2.51E-16 | TRUE | TRUE |

[a]SNPs with unadjusted p-value < 0.05 are shown in this table.

SNP: Variant identifier.

OR: Odds ratio.

SE: Standard error of odds ratio estimate.

UNADJ P: Unadjusted p-value.

BONF: Achieves significance via Bonferroni correction (TRUE/FALSE).

FDR: Achieves significance via FDR correction (TRUE/FALSE).

**This is a sample table of Supplemental Table 14. The complete table is included as a separate Excel file.**

**Supplemental Table 15. DNA sequences cloned into luciferase reporter plasmid.**

| Locus | Description of cloned sequence | Sequence coordinates (bp, b38) | Size | SNP | SNP position (bp, b38) |
|---|---|---|---|---|---|
| PAM | DNA sequence centered on rs76177300 and the associated T2D non-risk allele | chr5:102807541-102807740 | 200 | rs76177300 | chr5:102807607-102807607 |

| | | | | | |
|---|---|---|---|---|---|
| PAM | DNA sequence centered on rs76177300 and the associated T2D risk allele | chr5:102807541-102807740 | 200 | rs76177301 | chr5:102807607-102807607 |
| PAM | DNA sequence encompassing rs76177300 and nearby PAM eQTLs with the T2D non-risk haplotype | chr5:102807541-102808251 | 711 | rs76177300; rs37013; rs183662 | chr5:102807607-102807607; chr5:102807814-102808014; chr5:102807857-102808057 |
| PAM | DNA sequence encompassing rs76177300 and nearby PAM eQTLs with the T2D risk haplotype | chr5:102807541-102808251 | 711 | rs76177300; rs37013; rs183663 | chr5:102807607-102807607; chr5:102807814-102808014; chr5:102807857-102808058 |
| PAM | DNA sequence extending -249 upstream to +1 downstream of a *PAM* TSS[a] | chr5:102865623-102865872 | 250 | NA | NA |

[a]This DNA sequence spans an ENCODE promoter-like cCRE (EH38E2395186) and lies within an islet-specific HMR that overlaps a chromatin accessibility peak in islets (Varshney et al. 2017; Abascal et al. 2020).

**This is a sample table of Supplemental Table 15. The complete table is included as a separate Excel file.**

**Supplemental Table 16. Key resources table.**

| Critical commercial assays | Brand | Catalog number |
|---|---|---|
| EZ DNA Methylation-Gold Kit | Zymo Research | Cat# D5005 |
| Wizard Genomic DNA Purification Kit | Promega | Cat# A1120 |
| Genomic DNA Clean & Concentrator-25 | Zymo Research | Cat# D4065 |
| Genomic DNA Reagents and ScreenTape | Zymo Research | Cat# 5067-5366, 5067-5365 |
| DNA Clean & Concentrate-5 | Zymo Research | Cat# D4004 |
| D5000 Reagents and ScreenTape | Agilent | Cat# 5067-5589, 5067-5588 |
| 2x KAPA HiFi HotStart Uracil+ ReadyMix | Roche | Cat# 07959052001 |

**This is a sample table of Supplemental Table 16. The complete table with all resources used is included as a separate Excel file.**

### SUPPLEMENTAL FIGURE LEGENDS

**Figure S1. Characteristics of individual and reference islet HMRs from non-T2D donors. (A)** Density plot of HMR lengths by donor. The x-axis of the plot is visually limited to the range of 0 to 5000 bp for visibility. The red and black lines denote the median and mean HMR lengths, respectively. **(B)** Density plot of CpG count at HMRs by donor. The x-axis of the plot is visually limited to the 0 to 85 range for visibility. The red and black lines denote the median and mean CpG counts, respectively. **(C)** Box plots showing methylation levels at HMRs by donor. Mean methylation levels at HMRs were calculated by averaging CpG methylation scores using the *bedtools map* function with option '-o = mean'. **(D)** Density plot of reference islet HMR lengths. The x-axis of the plot is visually limited to the range of 0 to 7500 bp for visibility. The red and black lines denote the median and mean lengths, respectively. **(E)** Density plot of symmetric CpG counts at reference islet HMRs. These counts only represent symmetric CpG sites measured across all four non-T2D donors (n = 29,954,671). The x-axis of the plot is visually limited to the range of 0 to 450 for visibility. The red and black lines denote the median and mean CpG counts, respectively. **(F)** Box plot showing methylation levels at reference islet HMRs. The reference CpG methylation profile for islets (see **Methods**) was used as input to the *bedtools map* function with option '-o = mean' to compute the average methylation score for each HMR. **(G)** Heatmap depicting pairwise correlations across donors based on average methylation levels at non-reference islet HMRs. Spearman correlation coefficients are indicated by the color scale, and clusters were generated based on these values using complete linkage. H1 ESC and B cell WGBS datasets are included as outgroups to contextualize the strength of pairwise correlations between islet donors.

**Figure S2. Methylation differences at HMRs across islets and other diverse cell types captures their biological relationships. (A)** Dendrogram depicting hierarchical clustering of cell types based on methylation levels across a consensus HMR set. The matrix used to generate the *k*-means clustering heatmap in **Fig. 2A** was used as input to the *dist()* R function to build a distance matrix using the euclidean distance method, followed by hierarchical clustering using the *hclust()* R function with the complete linkage method. **(B)** Dot plot of elbow method to determine optimal number of clusters in *k*-means clustering for methylation heatmap. Total within-cluster sum of squares (WCSS) estimates are plotted on the y-axis against *k* values ranging from 1 to 15 on the x-axis. WCSS estimates were derived from the *kmeans()* function in R. **(C)** Heatmap of top representative TFs for *k*-means clusters represented in **Fig. 2A**. Scaled by row, color represents the fold change between the percentage of target regions containing a given motif relative to randomly selected background regions generated by the *HOMER* software.

**Figure S3. Top enriched traits in islets and other cell types group into cell-relevant clinical phenotypes.** Trait enrichment heatmap comparing SNP-trait associations in cell-type specific HMR datasets to those in the NHGR-EBI GWAS catalog. Scaled by row, color represents the fold change of a trait in a cell-type specific HMR dataset relative to its frequency in the catalog. *P*-values were calculated using a hypergeometric test: *p*-value < 0.05 (*), < 0.01 (**), < 0.001 (***).

**Figure S4. Phenotypic associations of T2D signals at islet-specific HMRs implicate pleiotropic effects extending beyond endocrine traits**. **(A)** Manhattan plot of phenotypic associations for 163 SNPs (MAF > 1%) with T2D association signals measured in either or both DIAMANTE and TDGGI studies and are found within islet-specific HMRs. The combined phenotypic associations for all SNPs are grouped and color-coded by phenotype group (x-axis) and -$\log_{10}$($p$-value) (y-axis). The dashed red line represents the per-SNP Bonferroni-corrected significance threshold for the minimum number of phenotypes tested per SNP after excluding associations with missing $p$-values ($p$-value < 0.05/1712). Across all SNPs, the number of phenotypes tested ranged from 1712 to 1714. The solid red line marks the global (i.e., experiment-wide) Bonferroni-corrected significance threshold calculated based on the total number of phenotype tests performed across all SNPs with non-missing $p$-values ($p$-value < 0.05/279346). SNP-phenotype associations that passed FDR correction on a per SNP basis are labeled on the plot and included among statistically significant phenotype associations. Nearest-neighbor genes of the islet-specific HMRs (see **Methods**) containing these signals are listed in the key and grouped by phenotype group. Genes appearing in multiple categories indicate phenotypic associations across more than one phenotype group. **(B)** Pie chart summarizing SNP-phenotype associations that met or exceeded per-SNP FDR correction. In total, 59 statistically significant association signals were identified across 22 clinical phenotypes organized into 11 broader phenotype groups. **(C)** Pie chart summarizing SNP-phenotype associations found at the *HLA-DRB5* locus. Among the significant SNP-phenotype associations shown in panel B, 37 are located within the MHC class II gene, *HLA-DRB5*. These hits span 10 clinical phenotypes that fall within the endocrine/metabolic, dermatologic, or neurological phenotype groups. **(D)** Pie chart summarizing SNP-phenotype associations found beyond the *HLA-DRB5* locus. After excluding associations at *HLA-DRB5* depicted in panel C, the remaining 22 map to 11 unique genes and are linked to 14 clinical phenotypes that group into 9 broader phenotype groups.

**Figure S5. Laboratory-trait associations at T2D-associated islet-specific HMRs implicate pleiotropic effects extending beyond key metabolic biomarkers in T2D. (A)** Manhattan plot of laboratory-trait associations for 163 SNPs (MAF > 1%) with T2D association signals measured in either or both DIAMANTE and T2DGGI studies and are found within islet-specific HMRs. Data points are grouped and color-coded by laboratory-test group on the x-axis, with corresponding -$\log_{10}$($p$-values) on the y-axis. The red line represents the global Bonferroni-corrected significance threshold ($p$-value < $9.96 \times 10^{-9}$) for 50,204 tests (308 labs per SNP). The black line indicates the FDR threshold ($p$-value < $6.38 \times 10^{-5}$). Laboratory-trait association signals that met or exceeded FDR are labeled on the plot. Nearest-neighbor genes of the islet-specific HMRs (see **Methods**) containing these statistically significant signals are listed in the key and grouped by clinical laboratory group. Genes appearing in multiple categories indicate laboratory-trait associations across more than one clinical laboratory group. **(B)** Pie chart summarizing SNP-lab associations that met or exceeded the FDR threshold. A total of 62 statistically significant associational signals were identified across 17 clinical-lab tests, which group into 4 broader clinical laboratory test groups; AC = automated count, M/V = Mass/Volume.

**Figure S6. Standard GWAS for T2D does not detect *PAM* locus signals observed in an HMR-WAS. (A)** QQ-plot from T2D HMR-WAS in BioVU subjects of European descent. The y-axis compares the observed -$\log_{10}$($p$-values) for T2D-SNP associations against their expected -$\log_{10}$($p$-values) under the null hypothesis of no association on the x-axis (in red). The *y=x* line in grey indicates the theoretical distribution where the observed and expected $p$-values are identical. The yellow lines represent a 95% confidence interval surrounding the theoretical distribution. The $\lambda$ value represents the genomic inflation factor. **(B)** Manhattan plot of GWAS results for T2D.

Markers with MAF > 1% on the Illumina $MEGA^{EX}$ array were tested for association with the T2D phecode (250.2). The blue and red lines represent the nominal (*p*-value = 0.05) and Bonferroni-corrected thresholds for significance (*p*-value < $9.05 \times 10^{-9}$), respectively. The black line marks the genome-wide significance threshold (*p*-value < $5.00 \times 10^{-8}$). The x-axis shows SNP genomic positions organized by chromosome, and the y-axis corresponds to the $-\log_{10}$(*p*-values) for their association with T2D. On chromosome 5, the purple data points denote an LD block of 30 SNPs at the *PAM* locus that achieved statistical significance for T2D association in our HMR-WAS, but not in a conventional GWAS (see **Fig. 2B**). On chromosome 10, the navy data points represent an LD block of 64 SNPs at the *TCF7L2* locus with statistically significant T2D association signals after FDR correction (*p*-value < $3.26 \times 10^{-7}$). **(C)** QQ plot from T2D GWAS in BioVU subjects of European descent. Refer to panel A for plot details.

**Figure S7. Chromatin profiling across cell types supports islet-specific enhancer activity at *PAM*-, *GCK*-, and *SLC2A2*-associated HMRs. (A)** Chromatin state alignment across islets and other WGBS-profiled cell types at the *PAM* locus. The *PAM*-associated islet-specific HMR (dashed box) displays a chromatin signature indicative of active enhancer activity found only in islets. The islet WGBS track shows mean methylation levels at non-mutated CpG sites across islet donors. ATAC-seq data for islets ("Islet 1" sample) and the other cell types were all obtained from Varshney et al. 2017. **(B)** Box plot comparing transcriptional activity of a *PAM* TSS-spanning sequence to an empty-vector control in INS1E and MIN6 β-cells (n = 6 per cell line). **(C)** Chromatin state alignment across islets and other WGBS-profiled cell types at the *GCK* locus. The *GCK*-associated islet-specific HMR (dashed box) exhibits an active enhancer chromatin state uniquely found in islets. **(D)** Box plot comparing transcriptional activity of a *GCK* TSS-spanning sequence to an empty-vector control (INS1E: n = 6 per group; MIN6: n = 4 for empty-vector, n = 6 for *GCK* TSS-spanning sequence). **(E)** Chromatin state alignment across islets and other WGBS-profiled cell types at the *SLC2A2* locus. Only islets show an active enhancer chromatin state at the *SLC2A2*-associated islet-specific HMR (dashed box). **(F)** Box plot comparing transcriptional activity of a *SLC2A2* TSS-spanning sequence to an empty-vector control (INS1E: n = 5 for empty-vector, n = 6 for *SLC2A2* TSS-spanning sequence; MIN6: n = 6 per group); Refer to panel A for track details shown in panels C and E. Fold-change in Firefly:*Renilla* luciferase activity relative to an empty-vector control is shown on the y-axis for panels B, D, and F. Significance was determined using two-sided unpaired *t*-tests: *p*-value < 0.05 (*), < 0.01 (**), < 0.001 (***).

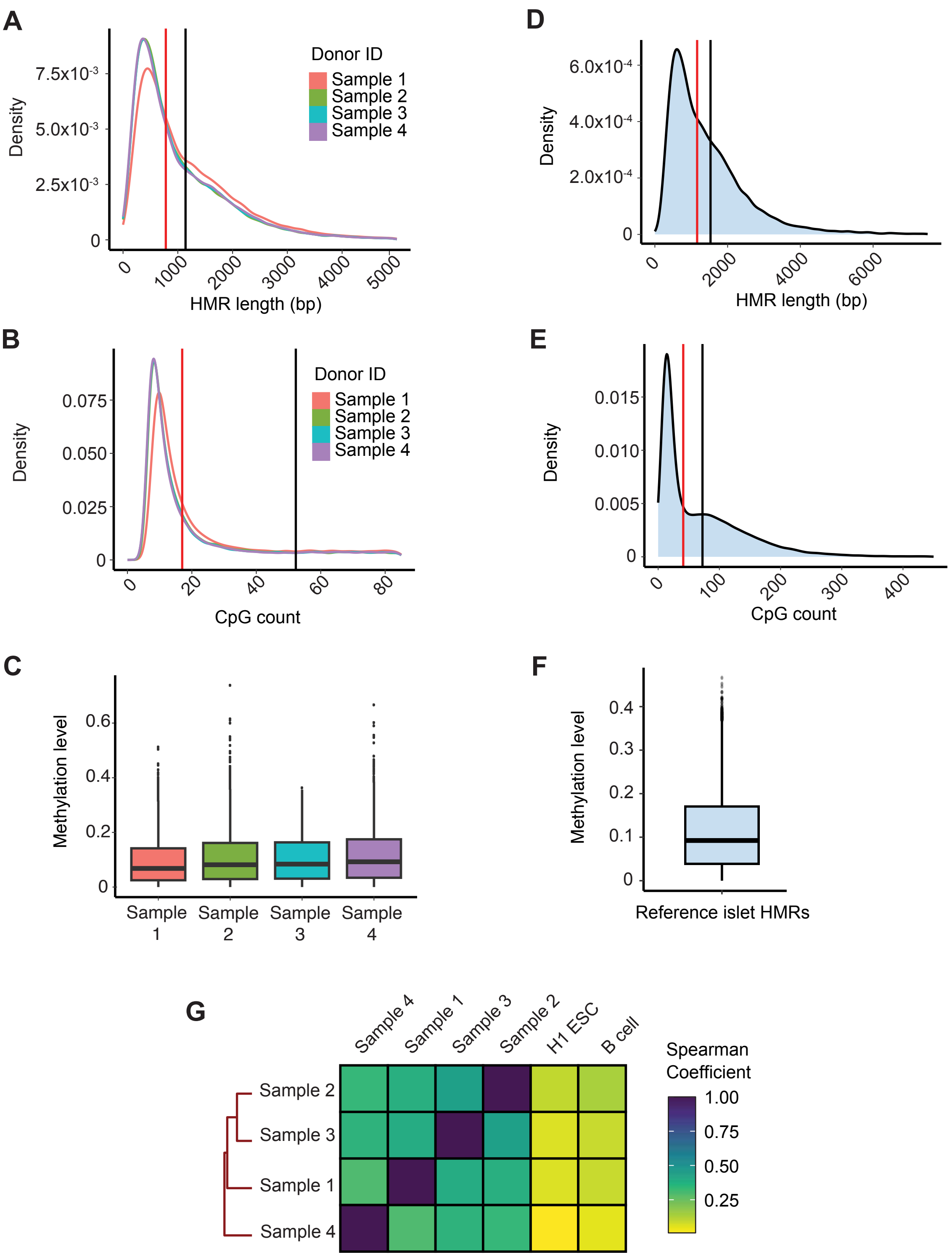

**Figure S1. Characteristics of individual and reference islet HMRs from non-T2D donors.** (A) Density plot of HMR lengths by donor. The x-axis of the plot is visually limited to the range of 0 to 5000 bp for visibility. The red and black lines denote the median and mean HMR lengths, respectively. (B) Density plot of CpG count at HMRs by donor. The x-axis of the plot is visually limited to the 0 to 85 range for visibility. The red and black lines denote the median and mean CpG counts, respectively. (C) Box plots showing methylation levels at HMRs by donor. Mean methylation levels at HMRs were calculated by averaging CpG methylation scores using the *bedtools map* function with option '-o = mean'. (D) Density plot of reference islet HMR lengths. The x-axis of the plot is visually limited to the range of 0 to 7500 bp for visibility. The red and black lines denote the median and mean lengths, respectively. (E) Density plot of symmetric CpG counts at reference islet HMRs. These counts only represent symmetric CpG sites measured across all four non-T2D donors (n= 29,954,671). The x-axis of the plot is visually limited to the range of 0 to 450 for visibility. The red and black lines denote the median and mean CpG counts, respectively. (F) Box plot showing methylation levels at reference islet HMRs. The reference CpG methylation profile for islets (see Methods) was used as input to the *bedtools map* function with option '-o = mean' to compute the average methylation score for each HMR. (G) Heatmap depicting pairwise correlations across donors based on average methylation levels at non-reference islet HMRs. Spearman correlation coefficients are indicated by the color scale, and clusters were generated based on these values using complete linkage. H1 ESC and B cell WGBS datasets are included as outgroups to contextualize the strength of pairwise correlations between islet donors.

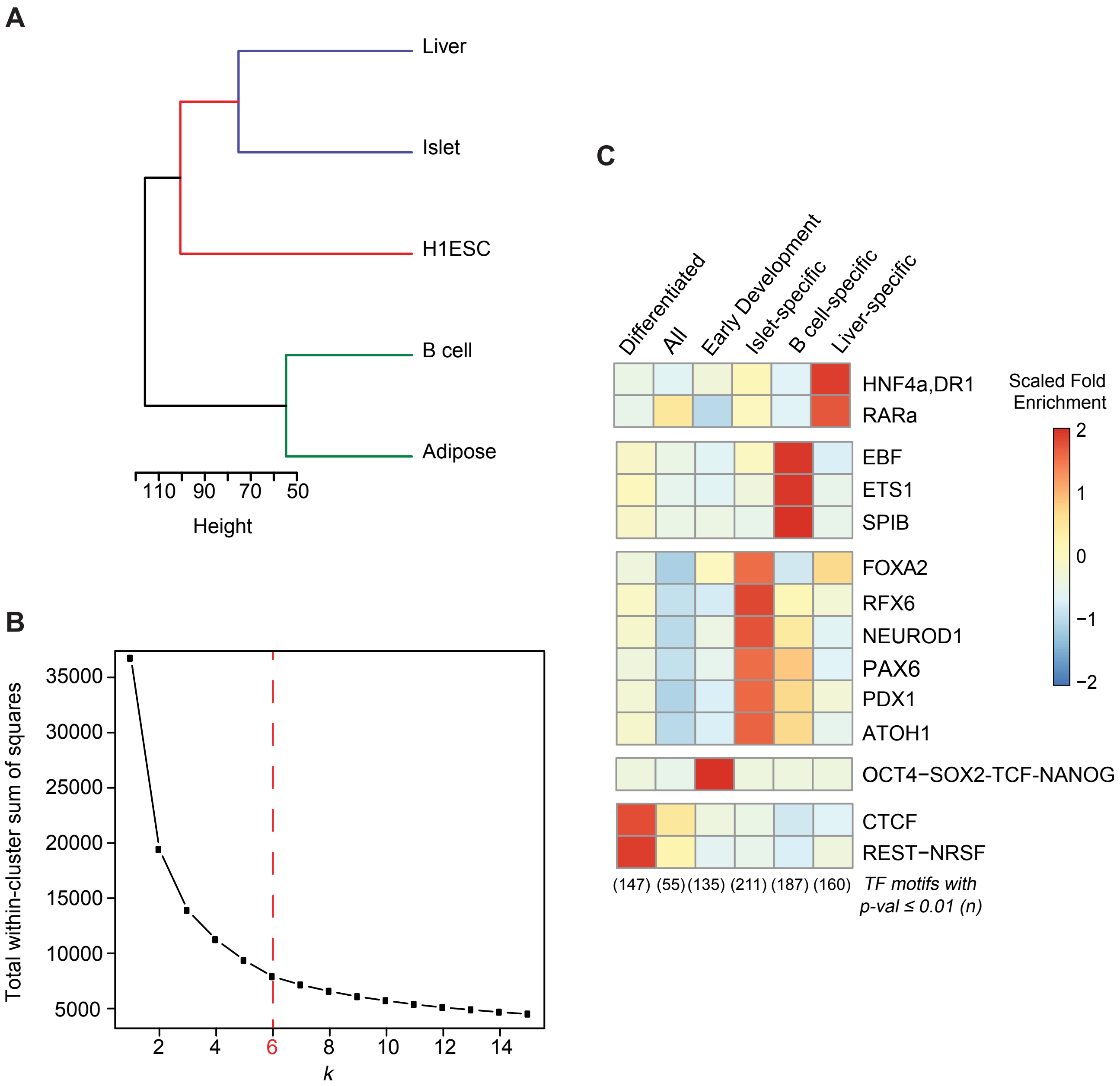

**Figure S2. Methylation differences at HMRs across islets and other diverse cell types captures their biological relationships.** (A) Dendrogram depicting hierarchical clustering of cell types based on methylation levels across a consensus HMR set. The matrix used to generate the *k*-means clustering heatmap in **Fig. 2A** was used as input to the *dist()* R function to build a distance matrix using the euclidean distance method, followed by hierarchical clustering using the *hclust()* R function with the complete linkage method. (B) Dot plot of elbow method to determine optimal number of clusters in *k*-means clustering for methylation heatmap. Total within-cluster sum of squares (WCSS) estimates are plotted on the y-axis against *k* values ranging from 1 to 15 on the x-axis. WCSS estimates were derived from the *kmeans()* function in R. (C) Heatmap of top representative TFs for *k*-means clusters represented in **Fig. 2A**. Scaled by row, color represents the fold-change between the percentage of target regions containing a given motif relative to randomly selected background regions generated by the HOMER software.

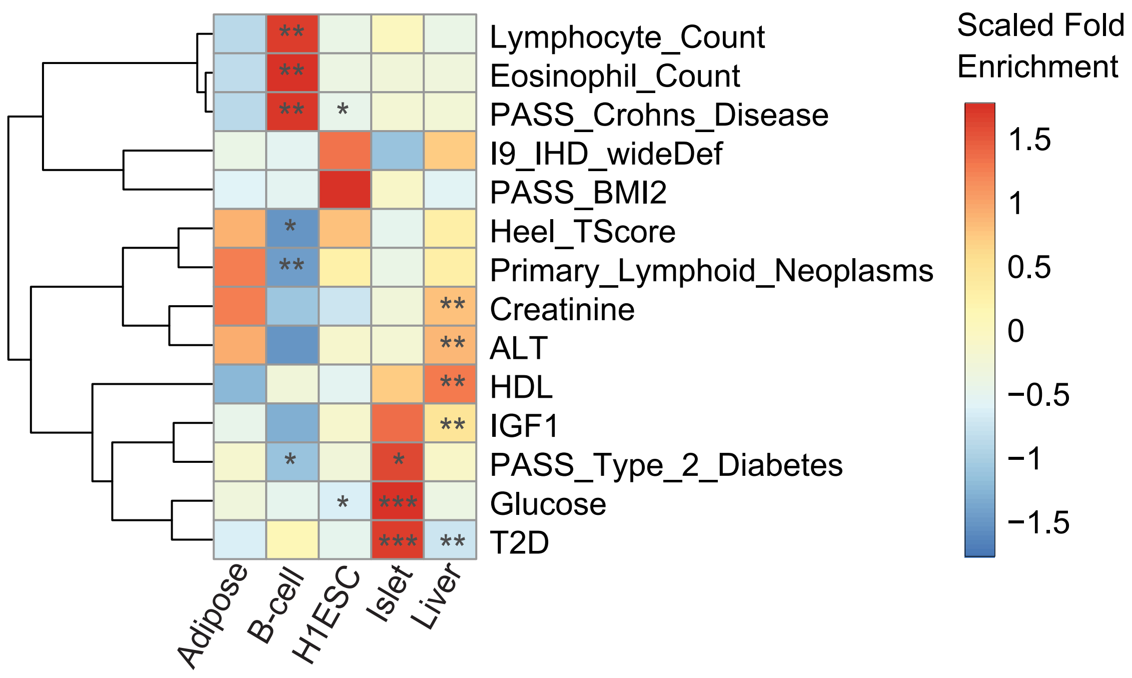

**Figure S3. Top enriched traits in islets and other cell types group into cell-relevant clinical phenotypes.** Trait enrichment heatmap comparing SNP-trait associations in cell-type specific HMR datasets to those in the NHGR-EBI GWAS catalog. Scaled by row, color represents the fold-change of a trait in a cell-type specific HMR dataset relative to its frequency in the catalog. *P*-values were calculated using a hypergeometric test: *p*-value < 0.05 (*), < 0.01 (**), < 0.001 (***).

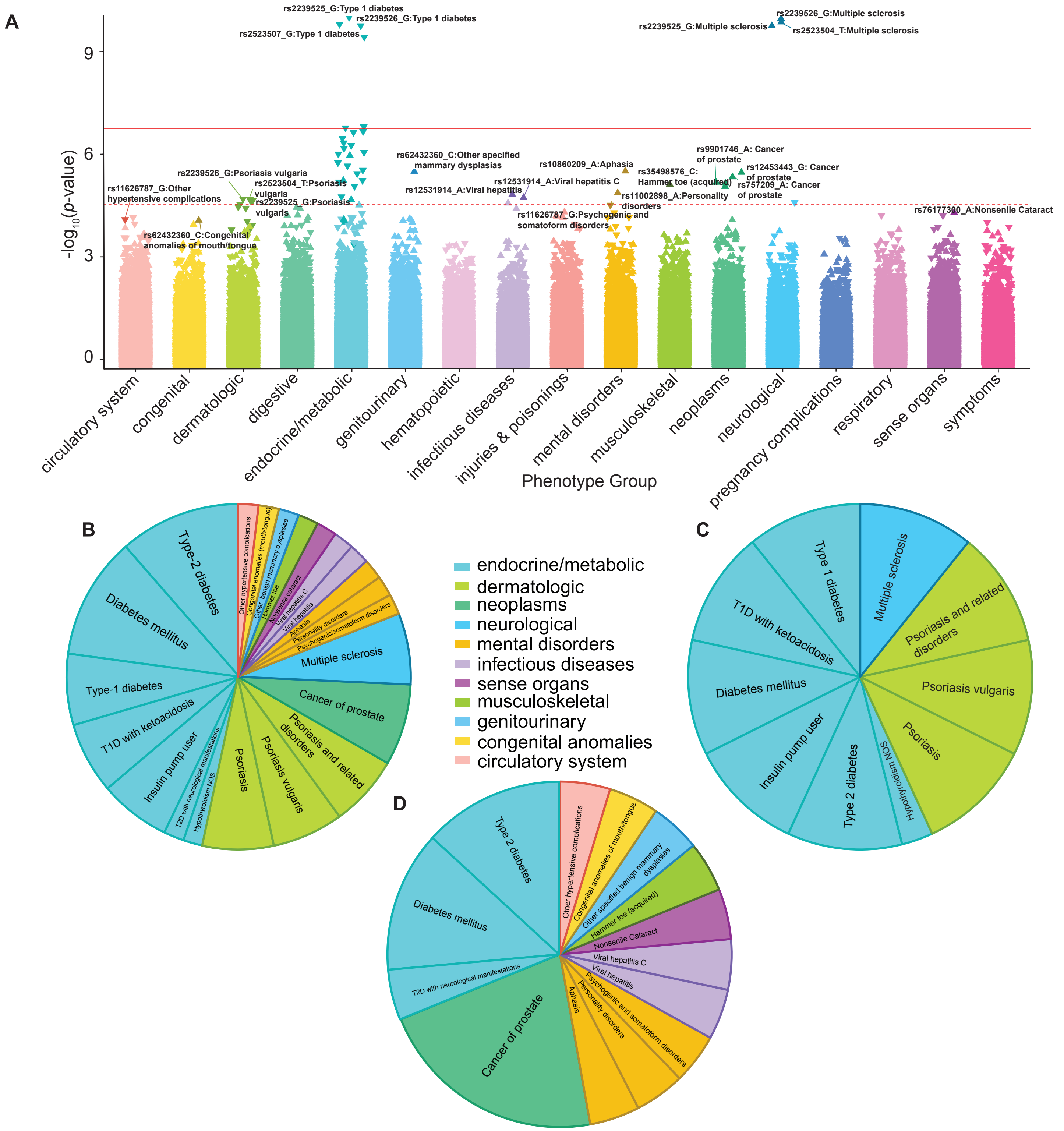

**Figures S4. Phenotypic associations of T2D signals at islet-specific HMRs implicate pleiotropic effects extending beyond endocrine traits.** (A) Manhattan plot of phenotypic associations for 163 SNPs (MAF > 1%) with T2D association signals measured in either or both DIAMANTE and TDGGI studies and are found within islet-specific HMRs. The combined phenotypic associations for all SNPs are grouped and color-coded by phenotype group (x-axis) and -$\log_{10}$(*p*-value) (y-axis). The dashed red line represents the per-SNP Bonferroni-corrected significance threshold for the minimum number of phenotypes tested per SNP after excluding associations with missing *p*-values (*p*-value < 0.05/1712). Across all SNPs, the number of phenotypes tested ranged from 1712 to 1714. The solid red line marks the global (i.e., experiment-wide) Bonferroni-corrected significance threshold calculated based on the total number of phenotype tests performed across all SNPs with non-missing p-values (*p*-value < 0.05/279346). SNP-phenotype associations that passed FDR correction on a per SNP basis are labeled on the plot and included among statistically significant phenotype associations. Nearest-neighbor genes of the islet-specific HMRs (see *Methods*) containing these signals are listed in the key and grouped by phenotype group. Genes appearing in multiple categories indicate phenotypic associations across more than one phenotype group. (B) Pie chart summarizing SNP-phenotype associations that met or exceeded per-SNP FDR correction. In total, 59 statistically significant association signals were identified across 22 clinical phenotypes organized into 11 broader phenotype groups. (C) Pie chart summarizing SNP-phenotype associations found at the *HLA-DRB5* locus. Among the significant SNP-phenotype associations shown in panel B, 37 are located within the MHC class II gene, *HLA-DRB5*. These hits span 10 clinical phenotypes that fall within the endocrine/metabolic, dermatologic, or neurological phenotype groups. (D) Pie chart summarizing SNP-phenotype associations found beyond the *HLA-DRB5* locus. After excluding associations at HLA-DRB5 depicted in panel C, the remaining 22 map to 11 unique genes and are linked to 14 clinical phenotypes that group into 9 broader phenotype groups.

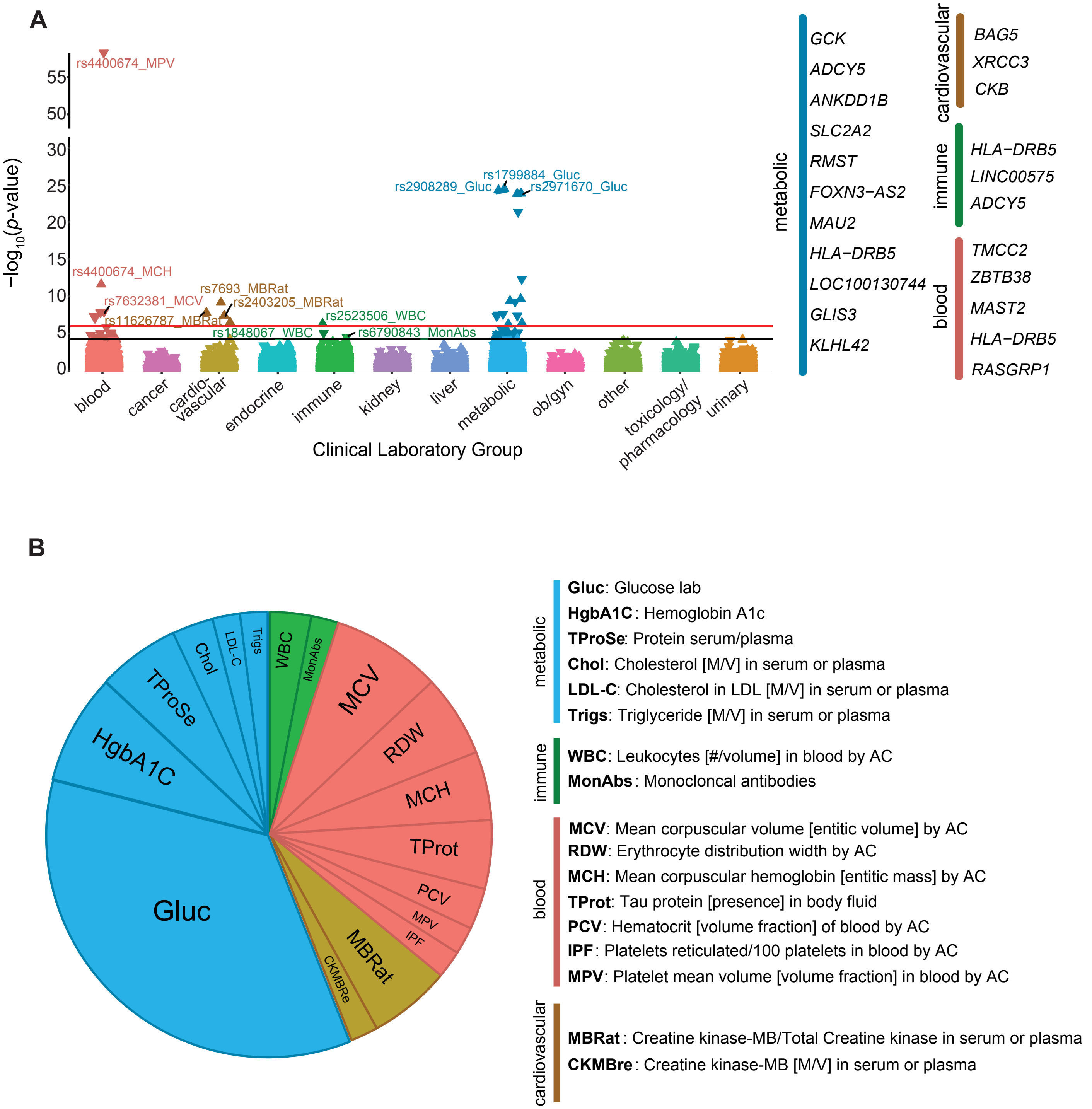

**Figure S5. Laboratory-trait associations at T2D-associated islet-specific HMRs implicate pleiotropic effects extending beyond key metabolic biomarkers in T2D.** (A) Manhattan plot of laboratory-trait associations for 163 SNPs (MAF > 1%) with T2D association signals measured in either or both DIAMANTE and T2DGGI studies and are found within islet-specific HMRs. Data points are grouped and color-coded by laboratory-test group on the x-axis, with corresponding $-\log_{10}$($p$-values) on the y-axis. The red line represents the global Bonferroni-corrected significance threshold ($p$-value < $9.96 \times 10^{-9}$) for 50,204 tests (308 labs per SNP). The black line indicates the FDR threshold ($p$-value < $6.38 \times 10^{-5}$). Laboratory-trait association signals that met or exceeded FDR are labeled on the plot. Nearest-neighbor genes of the islet-specific HMRs (see Methods) containing these statistically significant signals are listed in the key and grouped by clinical laboratory group. Genes appearing in multiple categories indicate laboratory-trait associations across more than one clinical laboratory group. (B) Pie chart summarizing SNP-lab associations that met or exceeded the FDR threshold. A total of 62 statistically significant associational signals were identified across 17 clinical-lab tests, which group into 4 broader clinical laboratory test groups; AC = automated count, M/V = Mass/Volume.

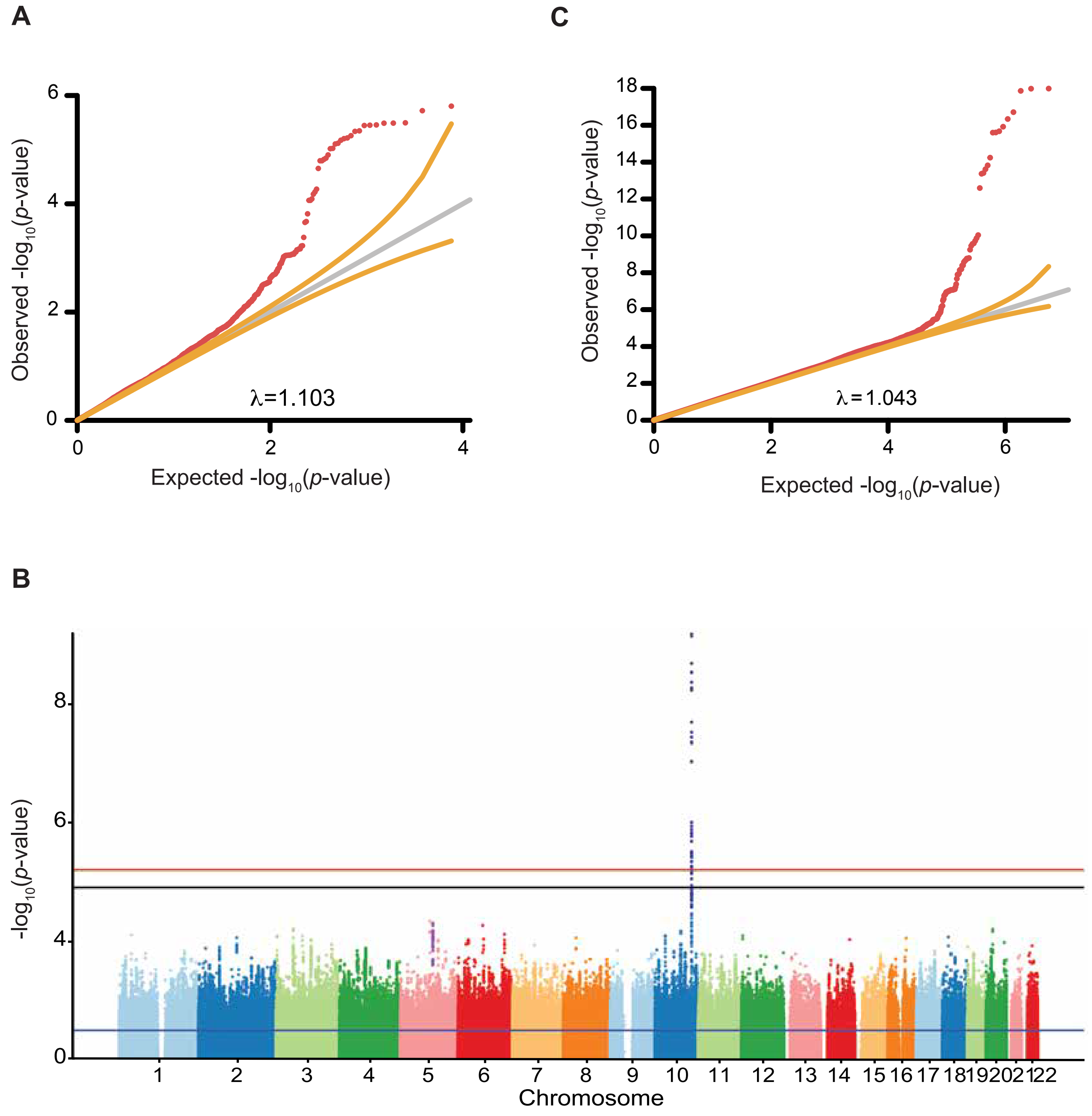

**Figure S6. Standard GWAS for T2D does not detect *PAM* locus signals observed in an HMR-WAS.** (A) QQ-plot from T2D HMR-WAS in BioVU subjects of European descent. The y-axis compares the observed $-\log_{10}$($p$-values) for T2D-SNP associations against their expected $-\log_{10}$($p$-values) under the null hypothesis of no association on the x-axis (in red). The y=x line in grey indicates the theoretical distribution where the observed and expected $p$-values are identical. The yellow lines represent a 95% confidence interval surrounding the theoretical distribution. The λ value represents the genomic inflation factor. (B) Manhattan plot of GWAS results for T2D. Markers with MAF > 1% on the Illumina MEGA$^{EX}$ array were tested for association with the T2D phecode (250.2). The blue and red lines represent the nominal ($p$-value = 0.05) and Bonferroni-corrected thresholds for significance ($p$-value < $9.05 \times 10^{-9}$), respectively. The black line marks the genome-wide significance threshold ($p$-value < $5.00 \times 10^{-8}$). The x-axis shows SNP genomic positions organized by chromosome, and the y-axis corresponds to the $-\log_{10}$($p$-values) for their association with T2D. On chromosome 5, the purple data points denote an LD block of 30 SNPs at the *PAM* locus that achieved statistical significance for T2D association in our HMR-WAS, but not in a conventional GWAS (see **Fig. 2B**). On chromosome 10, the navy data points represent an LD block of 64 SNPs at the *TCF7L2* locus with statistically significant T2D association signals after FDR correction ($p$-value < $3.26 \times 10^{-7}$). (C) QQ plot from T2D GWAS in BioVU subjects of European descent. Refer to panel A for plot details.

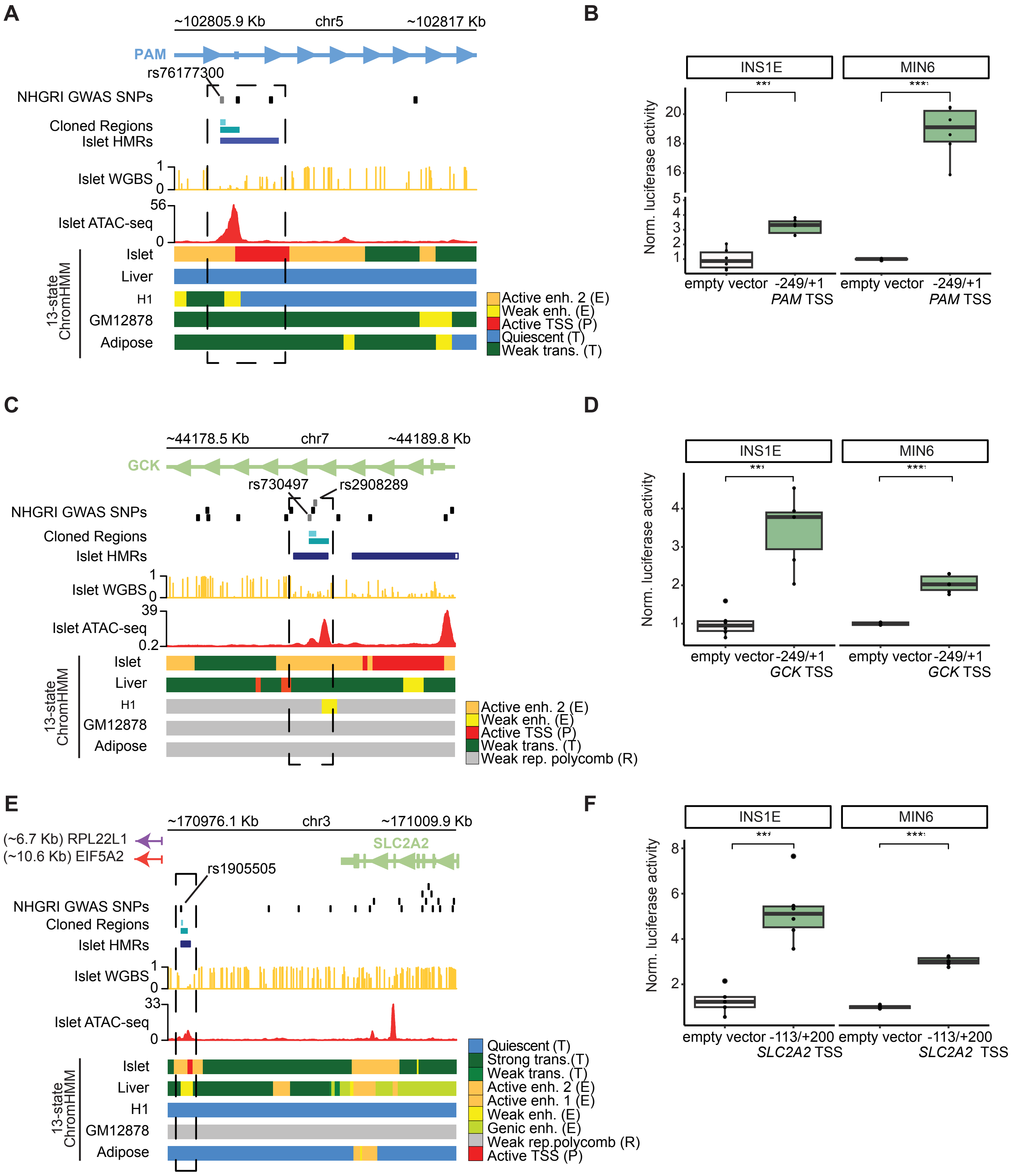

**Figure S7. Chromatin profiling across cell types supports islet-specific enhancer activity at *PAM*-, *GCK*-, and *SLC2A2*-associated HMRs.** (A) Chromatin state alignment across islets and other WGBS-profiled cell types at the *PAM* locus. The *PAM*-associated islet-specific HMR (dashed box) displays a chromatin signature indicative of active enhancer activity found only in islets. The islet WGBS track shows mean methylation levels at non-mutated CpG sites across islet donors. ATAC-seq data for islets ("Islet 1" sample) and the other cell types were all obtained from Varshney et al. 2017. (B) Box plot comparing transcriptional activity of a *PAM* TSS-spanning sequence to an empty-vector control in INS1E and MIN6 β-cells (n = 6 per cell line). (C) Chromatin state alignment across islets and other WGBS-profiled cell types at the *GCK* locus. The *GCK*-associated islet-specific HMR (dashed box) exhibits an active enhancer chromatin state uniquely found in islets. (D) Box plot comparing transcriptional activity of a *GCK* TSS-spanning sequence to an empty-vector control (INS1E: n = 6 per group; MIN6: n = 4 for empty-vector, n = 6 for *GCK* TSS-spanning sequence). (E) Chromatin state alignment across islets and other WGBS-profiled cell types at the *SLC2A2* locus. Only islets show an active enhancer chromatin state at the *SLC2A2*-associated islet-specific HMR (dashed box). (F) Box plot comparing transcriptional activity of a *SLC2A2* TSS-spanning sequence to an empty-vector control (INS1E: n = 5 for empty-vector, n = 6 for *SLC2A2* TSS-spanning sequence; MIN6: n = 6 per group); Refer to panel A for track details shown in panels C and E. Fold-change in Firefly:Renilla luciferase activity relative to an empty-vector control is shown on the y-axis for panels B, D, and F. Significance was determined using two-sided unpaired t-tests: *p*-value < 0.05 (*), < 0.01 (**), < 0.001 (***).
